# Automatic Three-Dimensional Cephalometric Landmarking via Deep Learning

**DOI:** 10.1101/2022.01.28.22269989

**Authors:** Gauthier Dot, Thomas Schouman, Shaole Chang, Frédéric Rafflenbeul, Adeline Kerbrat, Philippe Rouch, Laurent Gajny

## Abstract

The increasing use of three-dimensional (3D) imaging by orthodontists and maxillofacial surgeons to assess complex dentofacial deformities and plan orthognathic surgeries implies a critical need for 3D cephalometric analysis. Although promising methods were suggested to localize 3D landmarks automatically, concerns about robustness and generalizability restrain their clinical use. Consequently, highly trained operators remain needed to perform manual landmarking. In this retrospective diagnostic study, we aimed to train and evaluate a deep learning (DL) pipeline based on SpatialConfiguration-Net for automatic localization of 3D cephalometric landmarks on computed tomography (CT) scans. A retrospective sample of consecutive presurgical CT scans was randomly distributed between a training/validation set (*n* = 160) and a test set (*n* = 38). The reference data consisted in 33 landmarks, manually localized once by 1 operator (*n* = 178) or twice by 3 operators (*n* = 20, test set only). After inference on the test set, one CT scan showed “very low” confidence level predictions; we excluded it from the overall analysis but still assessed and discussed the corresponding results. The model performance was evaluated by comparing the predictions with the reference data; the outcome set included localization accuracy, cephalometric measurements and comparison to manual landmarking reproducibility. On the hold-out test set, the mean localization error was 1.0 ± 1.3mm, while success detection rates for 2.0, 2.5 and 3.0mm were 90.4%, 93.6% and 95.4%, respectively. Mean errors were −0.3 ± 1.3° and −0.1 ± 0.7mm for angular and linear measurements, respectively. When compared to manual reproducibility, the measurements were within the Bland-Altman 95% limits of agreement for 91.9% and 71.8% of skeletal and dentoalveolar variables, respectively. To conclude, while our DL method still requires improvement, it provided highly accurate 3D landmark localization on a challenging test set, with a reliability for skeletal evaluation on par with what clinicians obtain.

## Introduction

Three-dimensional (3D) computed tomography (CT) or cone beam CT (CBCT) scans are increasingly used by orthodontists and maxillofacial surgeons for diagnosis and treatment planning purposes. While two-dimensional (2D) radiographs are still sufficient for most of orthodontic patients, 3D scans allow clinicians to assess complex maxillomandibular deformities and craniofacial anomalies, improving diagnosis and treatment planning for those patients (American Academy of Oral and Maxillofacial Radiology 2013; Kapila and Nervina 2015). More specifically, 3D images are now widely used for the planning of computer-assisted orthognathic surgical procedures (Alkhayer et al. 2020). For each patient, this planning is usually performed by a technician, following a surgeon’s prescription based on clinical examination and cephalometric analysis of the 3D scans (Xia et al. 2009). Cephalometric analysis is used to measure the deviation of the skeletal and dentoalveolar parts of the maxilla and the mandible in relation to the skull base, using measurements between specific landmarks placed on each of these structures. The reference method for 3D cephalometric analysis is manual landmarking, which requires around 15 minutes for a highly experienced and trained operator (Hassan et al. 2013; Dot et al. 2021).

The automatization of 3D cephalometric landmarking has been an active research field over the last decade, as the clinical dissemination of such a method would decrease the burden of manual landmarking. Two systematic reviews recently reported on the accuracy of such automated methods (Dot et al. 2020; Schwendicke, Chaurasia, et al. 2021). Both yielded promising results for deep learning (DL) based methods, which outperformed previously proposed knowledge-based, atlas-based or shallow learning-based methods. DL methods published in the last few years can localize 3D cephalometric landmarks with great accuracy, often under the 2-mm threshold of clinical acceptability (Lee et al. 2019; O’Neil et al. 2019; Torosdagli et al. 2019; Lang et al. 2020; Ma et al. 2020; Yun et al. 2020; Zhang et al. 2020; Bermejo et al. 2021; Chen et al. 2021; Kang et al. 2021; Liu et al. 2021; Chen et al. 2022). The studies showing the best results usually formulate landmark detection as a regression problem, using landmark heatmap regression methods (Zhang et al. 2020; Chen et al. 2021). However, the evaluation of the published models is often limited to few landmarks, and both systematic reviews noted a high risk of bias in the reporting of these studies, mainly because the description of the database/reference was limited and because the accuracy scores were calculated from within-sample validation datasets or very small hold-out test sets (<10 scans). As a result, major concerns remain about the robustness and generalizability of DL methods for 3D cephalometric landmarking, highlighting the need for additional evaluation studies with clinically relevant datasets, clear reference data and broader outcome metrics (Dot et al. 2020; Schwendicke, Chaurasia, et al. 2021).

Recently, the fully convolutional neural network (CNN) SpatialConfiguration-Net (SCN) was proposed as a heatmap regression method integrating a spatial configuration module for landmark localization (Payer et al. 2019). SCN has shown impressive results for the localization of anatomic landmarks on datasets of hand radiographs, lateral cephalograms and spine CT scans, but has yet to be evaluated on craniomaxillofacial CT scans (Payer et al. 2019; Sekuboyina et al. 2021). One difficulty to overcome is data size, as high resolution Head CT scans exceed the memory capacity of a typical graphical processing unit (GPU). There are two solutions to overcome this obstacle: 1) downsampling the scans by decreasing their resolution; 2) implementing the CNNs on small 3D image patches. However, downsampled data necessarily result in less accurate landmark localization, while image patches oftentimes lack volumetric context. But a 2-step, coarse-to-fine approach combining both methods could overcome these limitations (Chen et al. 2021; Sekuboyina et al. 2021).

The main goal of this diagnostic accuracy study was to design and implement a coarse-to-fine DL method based on SCN for automatic landmark localization (the index test), before thoroughly comparing its diagnostic performance with respect to manual landmarking (the reference test) on a hold-out test dataset of craniomaxillofacial CT scans from clinical practice.

## Materials and Methods

This study was approved by an appropriate Institutional Review Board (IRB No. CRM-2001-051) and its reporting followed recently published recommendations on artificial intelligence in dental research (Schwendicke, Singh, et al. 2021).

### Dataset

Two hundred presurgical CT scans, randomly selected and anonymized, were obtained from a retrospective sample described in a previous study (Dot et al. 2022), consisting of consecutive patients having undergone orthognathic surgery between January 2017 and December 2019 in a single maxillofacial surgery department. Patients referred to this university hospital located in a cosmopolitan European capital city were ethnically diverse and presented a variety of dentofacial deformities within the scope of orthognathic surgery (maxilla and/or mandible surgery, usually performed along with orthodontic treatment). Two subjects refused to participate; their data was excluded from the dataset. 198 subjects (198 anonymized presurgical CT scans) were eventually included in our dataset and randomly distributed among a training set (*n* = 128), a validation set (*n* = 32) and a test set (*n* = 38) (Appendix Figure 1). The subjects had a mean age of 27 ± 11 years (minimum age 14, maximum age 60) and 58.6% were females (*n* = 116). 89.4% of the CT scans (*n* = 177) showed metallic artefacts (orthodontic materials, metallic dental fillings or crowns) and 95.4% of the CT scans (*n* = 189) were acquired on the same CT machine. The scans had an average of 744 slices with a mean in-plane pixel size of 0.45*0.45mm^2^, mean field of view of 229mm and mean slice thickness of 0.33mm. Full CT scans and patient characteristics are detailed in Appendix Table 1.

### Manual landmarking (Reference Test)

Thirty-three landmarks, divided into skeletal (*n* = 21) and dental (*n* = 12) landmarks (Fig. 1A), were manually annotated on each CT scan, either once (*n* = 178) by operator #1 (a trained orthodontist with 5 years of clinical experience), or twice (*n* = 20) by operators #1, #2 (a trained orthodontist with 5 years of clinical experience) and #3 (a final year postgraduate maxillofacial surgeon). The reference data used to train and test our DL model were either the single annotations (*n* = 178) or the average of the 6 annotations (*n* = 20, test set only). The scans annotated six times were part of a previous repeatability and reproducibility (R&R) study, which could be used to evaluate intra and interobserver variability of the reference test (Dot et al. 2021). In the test set, some CT scans showed missing dental landmarks: 16O (*n* = 1), 26O (*n* = 1), 31A (*n* = 2), 31E (*n* = 2), 36O (*n* = 1), 46O (*n* = 2). Landmark definitions and landmarking procedure are detailed in the appendix.

**Figure 1.**
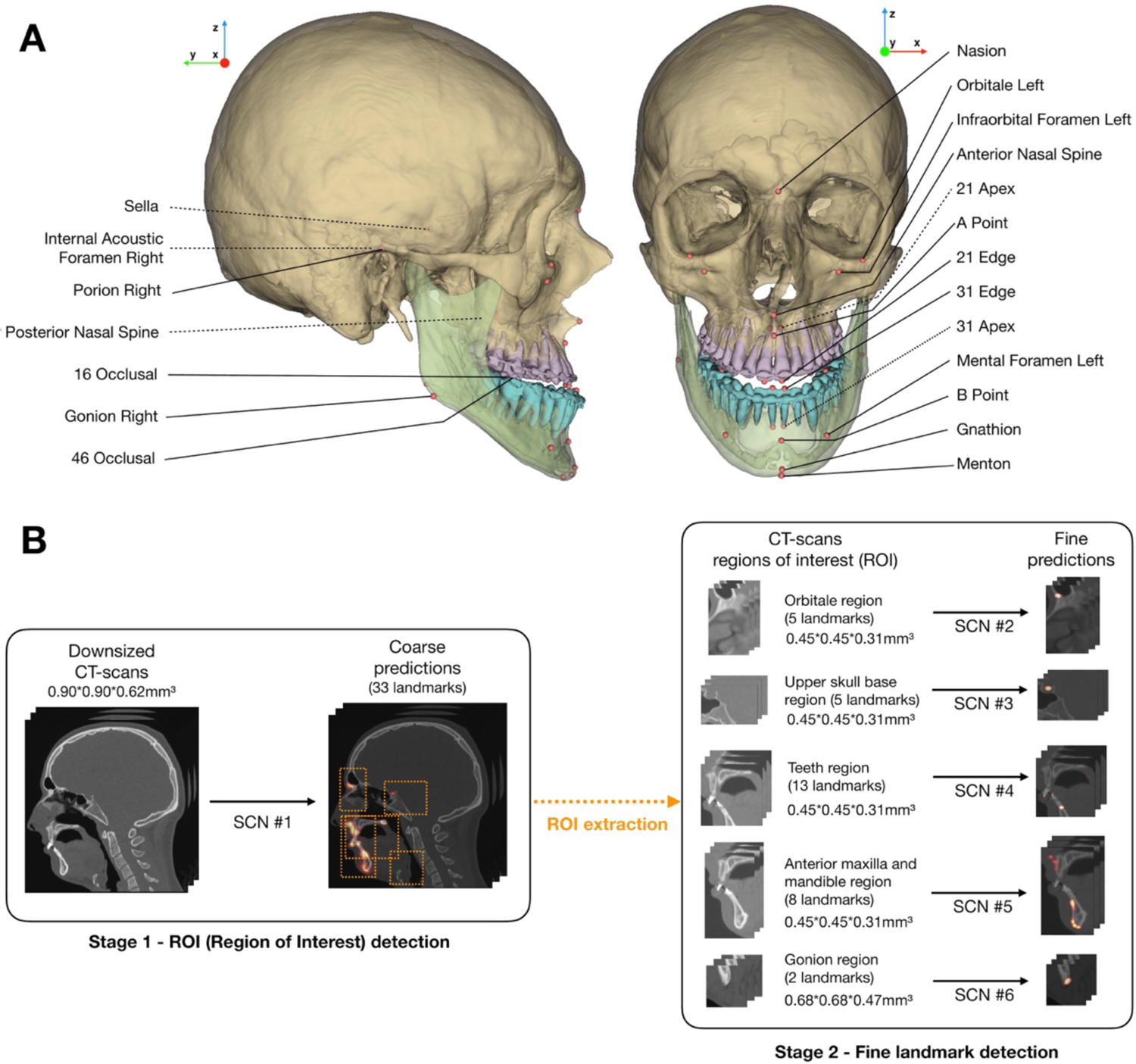
Landmarks and pipeline of the deep learning model. (**A**) Illustration of the set of 33 landmarks; bilateral landmarks are named once; dotted lines show landmarks localized inside the skull; (**B**) 2-stage method used for model inference. SCN, SpatialConfiguration-Net; ROI, region of interest.

### Deep learning-based landmarking (Index Test)

The DL model implemented in this study was the publicly available SCN described by Payer et al. (Payer et al. 2019), running in Tensorflow v1.15.0 on our laboratory workstation (CPU AMD Ryzen 9 3900X 12-Core; 128 Gb RAM; GPU Nvidia Titan RTX 24Gb). The pipeline used to train the network followed a coarse-to-fine approach: 1) to keep most of the volumetric context, we trained a first network (SCN#1) on downsampled-resolution full scans; 2) to localize the landmarks more accurately within selected regions of interest (ROIs), five networks (SCN#2 to SCN#6) were trained on selected full-resolution ROIs (Fig. 1B). The coordinates of each local heatmap maxima were considered as the predicted landmark positions. The confidence in a network prediction was evaluated as “very low” when the heatmap maximum value was below a threshold established from the validation results. Please refer to the appendix for additional implementation details.

Inference (prediction made by the trained model) was performed on our hold-out test set (*n* = 38) following a 2-stage method (Fig. 1B). At stage 1, SCN#1 predicted the “coarse” localization of the landmarks, which was then used to extract the 5 ROIs. At stage 2, SCN#2 to SCN#6 predicted the “fine” localization of the landmarks in each ROI along with the confidence in the prediction. This method systematically localized 33 landmarks for each CT scan. In CT scans with missing landmarks (*i*.*e*. missing teeth), the corresponding predictions were considered as missing values and deleted by the operator.

### Evaluation

If a CT scan showed several “very low” confidence levels in coordinate predictions, the subject was considered as an outlier case. To evaluate the overall localization performance on our test set, three commonly-used criteria were computed for each landmark (Wang et al. 2016): 1) mean radial error (MRE) – mean Euclidian distance between the reference landmark and the predicted landmark; 2) success detection rate (SDR) – proportion of landmarks located with radial errors under 2mm, 2.5mm, 3mm; 3) minimum and maximum radial error.

Conventional 2D cephalometric measurements (Appendix Table 4) were computed using orthogonal projections of the 3D landmarks on a midsagittal plane computed using a previously published automated method (Pinheiro et al. 2019). Additionally, the accuracy of Frankfort horizontal (FH) plane construction (porion right/left and orbitale left) was evaluated.

The predicted landmarks and cephalometric variables were compared to the Bland-Altman 95% limits of agreement (LoA) of manual landmarking and cephalometric measurement reproducibility, computed from a previous R&R study (Dot et al. 2021) following ISO norm 5725 (ISO 5725-2:2019). More details are provided in the appendix.

### Statistical analysis

Continuous variables were presented as means ± standard deviations; categorical variables were expressed as numbers and percentages. We first assessed the normality of the data using Shapiro-Wilk normality test, and then applied Wilcoxon and Student t-tests for nonparametric and parametric data, respectively; p-values < 0.05 were considered statistically significant.

## Results

### Training, testing and outlier case

Training time for one network on one GPU was about 48 hours and inference required around 1 minute per CT scan. One CT scan from a patient exhibiting cleidocranial dysplasia showed several predictions (A Point and several dental landmarks) with “very low” confidence levels. It was therefore considered as an outlier case and was excluded from the overall analysis, although individual localization performance was assessed and discussed.

### Localization performance

On our test set without the outlier case (*n* = 37), MRE for all landmarks was 1.0mm ± 1.3mm and SDRs for all landmarks were 90.4%, 93.6% and 95.4%, using 2mm, 2.5mm and 3mm precision ranges, respectively (Table 1). Thirteen landmarks (39.4%) showed SDRs at 2mm of 100%; 24 landmarks (72.7%) showed SDRs at 2mm over 90%, and 5 landmarks (15.2%) showed SDRs at 2mm under 80% (B point, gonion left and right, orbitale left and right). Additional results, including the outlier case and validation set evaluations, are reported in Appendix Tables 6, 7 and 8. When comparing scans with references constructed from 1 or 6 annotations, 3 landmarks exhibited statistically significantly larger errors when constructed from 6 annotations instead of 1 annotation: orbitale left, 11 incisal edge and 41 incisal edge.

**Table 1.** Mean radial errors (mm), success detection rates (% (*n*)) and minimum/maximum radial error (mm) for each landmark on the hold-out test set without the outlier case (*n* = 37). MRE, mean radial error; SD, standard deviation; Min., minimum radial error; Max., maximum radial error; L, left; R, right.

|  | MRE ± SD | <2mm | <2.5mm | <3mm | Min. | Max. |
| --- | --- | --- | --- | --- | --- | --- |
| 11 Apex | 0.7 ± 0.4 | 100 (37) | 100 (37) | 100 (37) | 0.2 | 1.5 |
| 11 Edge | 0.4 ± 0.3 | 100 (37) | 100 (37) | 100 (37) | 0.1 | 1.3 |
| 16 Occlusal | 1.3 ± 2.4 | 94.4 (34) | 94.4 (34) | 94.4 (34) | 0.1 | 11.2 |
| 21 Apex | 0.7 ± 0.3 | 100 (37) | 100 (37) | 100 (37) | 0.2 | 1.9 |
| 21 Edge | 0.5 ± 0.3 | 100 (37) | 100 (37) | 100 (37) | 0.1 | 1.4 |
| 26 Occlusal | 1.2 ± 2.4 | 94.4 (34) | 94.4 (34) | 94.4 (34) | 0.1 | 11.4 |
| 31 Apex | 0.9 ± 1.4 | 97.1 (34) | 97.1 (34) | 97.1 (34) | 0.2 | 8.7 |
| 31 Edge | 0.6 ± 1.1 | 94.4 (33) | 97.1 (34) | 97.1 (34) | 0.1 | 6.7 |
| 36 Occlusal | 1.5 ± 2.9 | 91.7 (33) | 91.7 (33) | 91.7 (33) | 0.2 | 11.3 |
| 41 Apex | 0.6 ± 0.3 | 100 (37) | 100 (37) | 100 (37) | 0.2 | 1.3 |
| 41 Edge | 0.5 ± 0.2 | 100 (37) | 100 (37) | 100 (37) | 0.1 | 1.3 |
| 46 Occlusal | 0.9 ± 1.8 | 97.2 (35) | 97.2 (35) | 97.2 (35) | 0.6 | 11.0 |
| A Point | 1.1 ± 0.9 | 89.2 (33) | 91.9 (34) | 91.9 (34) | 0.2 | 3.9 |
| Anterior Nasal Spine | 0.7 ± 0.7 | 94.6 (35) | 94.6 (35) | 97.3 (36) | 0.1 | 3.2 |
| B Point | 1.7 ± 1.5 | 67.6 (25) | 81.1 (30) | 91.9 (34) | 0.3 | 8.5 |
| Gnathion | 1.6 ± 0.6 | 91.9 (34) | 97.3 (36) | 100 (37) | 0.3 | 2.5 |
| Gonion L | 1.9 ± 1.7 | 70.3 (26) | 75.7 (28) | 86.5 (32) | 0.3 | 7.3 |
| Gonion R | 2.1 ± 1.4 | 48.7 (18) | 70.3 (26) | 73.0 (27) | 0.3 | 6.9 |
| Infraorbital Foramen L | 0.6 ± 0.3 | 100 (37) | 100 (37) | 100 (37) | 0.2 | 2.0 |
| Infraorbital Foramen R | 0.6 ± 0.5 | 97.3 (36) | 100 (37) | 100 (37) | 0.1 | 2.4 |
| Internal Acoustic Foramen L | 0.6 ± 0.4 | 100 (37) | 100 (37) | 100 (37) | 0.2 | 1.9 |
| Internal Acoustic Foramen R | 0.6 ± 0.6 | 97.3 (36) | 97.3 (36) | 97.3 (36) | 0.1 | 3.9 |
| Mental Foramen L | 0.4 ± 0.2 | 100 (37) | 100 (37) | 100 (37) | 0.1 | 0.8 |
| Mental Foramen R | 0.4 ± 0.3 | 100 (37) | 100 (37) | 100 (37) | 0.1 | 1.3 |
| Menton | 1.6 ± 0.6 | 94.6 (35) | 97.3 (36) | 100 (37) | 0.4 | 2.6 |
| Nasion | 0.6 ± 0.3 | 100 (37) | 100 (37) | 100 (37) | 0.1 | 1.9 |
| Orbitale L | 2.7 ± 2.0 | 43.2 (16) | 56.8 (21) | 67.6 (25) | 0.1 | 8.8 |
| Orbitale R | 2.6 ± 2.3 | 56.8 (21) | 67.6 (25) | 70.3 (26) | 0.3 | 9.7 |
| Pogonion | 1.1 ± 0.6 | 89.2 (33) | 97.3 (36) | 100 (37) | 0.2 | 3.0 |
| Porion L | 1.1 ± 0.5 | 89.2 (33) | 100 (37) | 100 (37) | 0.2 | 2.3 |
| Porion R | 1.3 ± 0.7 | 86.5 (32) | 89.2 (33) | 100 (37) | 0.3 | 2.8 |
| Posterior Nasal Spine | 0.5 ± 0.4 | 100 (37) | 100 (37) | 100 (37) | 0.1 | 1.5 |
| Sella | 0.8 ± 0.4 | 100 (37) | 100 (37) | 100 (37) | 0.2 | 2.0 |

### Cephalometric measurements

On our test set without the outlier case (*n* = 37), mean differences between the reference and predicted measurements were −0.3 ± 1.3° for angular observations and −0.1 ± 0.7mm for linear observations (Table 2). 96.7% (*n* = 322) of the skeletal measurements and 83.8% (*n* = 181) of the dentoalveolar measurements showed errors inferior to 2mm/2°. The mean absolute angular distance between predicted and reference FH planes was 0.4 ± 0.3°, and all the measurements were inferior to 2°.

**Table 2.** Mean errors (mm) and success detection rates (% (*n*)) for each cephalometric variable on the hold-out test set without the outlier case (*n* = 37). SD, standard deviation.

|  | Mean | SD | <2mm/2° |
| --- | --- | --- | --- |
| **Skeletal** |  |  |  |
| SNA (°) | -0.1 | 0.7 | 100 (37) |
| SNB (°) | -0.0 | 0.7 | 100 (37) |
| ANB (°) | -0.1 | 0.2 | 100 (37) |
| ANS-PNS / Go-Gn (°) | -0.1 | 1.3 | 91.9 (34) |
| S-Na / Go-Gn (°) | 0.0 | 1.4 | 83.8 (31) |
| Pog to NB (mm) | 0.0 | 0.4 | 100 (37) |
| A to MSP (mm) | 0.0 | 0.3 | 100 (37) |
| B to MSP (mm) | -0.3 | 0.6 | 97.3 (36) |
| Pog to MSP (mm) | -0.2 | 0.7 | 97.3 (36) |
| **Dentoalveolar** |  |  |  |
| SN / Occlusal plane (°) | -0.2 | 1.2 | 97.1 (34) |
| Upper inc / ANS-PNS (°) | -0.8 | 1.5 | 75.7 (28) |
| Upper inc to NA (mm) | 0.2 | 0.5 | 100 (37) |
| Inter-incisal angle (°) | -1.1 | 1.9 | 54.3 (19) |
| Lower inc / Go-Gn (°) | -0.4 | 1.8 | 77.1 (27) |
| Lower inc to NB (mm) | -0.1 | 1.0 | 97.3 (36) |

### Comparison with manual landmarking and measurement reproducibility

On our test set without the outlier case (*n* = 37), when comparing predicted landmark coordinates in the -*x*, -*y* and -*z* directions with manual landmarking repeatability, 90.7% (*n* = 2114) of the skeletal coordinates and 65.4% (*n* = 871) of the dental coordinates were within 95% LoA (Appendix Table 9). When comparing predicted cephalometric measurement errors with manual measurement repeatability, 91.9% (*n* = 306) of the skeletal variables and 71.8% (*n* = 155) of the dentoalveolar variables were within 95% LoA (Appendix Table 10). For the scans included in the R&R study (without the outlier case, *n* = 19), localization and measurement error boxplots for the manual and automatic methods are shown in Figure 2. Bland-Altman plots showing the deviations of manual and automatic landmarking localizations and cephalometric measurements for the scans included in the R&R study are reported in the Appendix. Automatic localization errors were statistically significantly larger than manual landmarking errors (Fig. 2) for 5 skeletal landmarks (32.8%), 10 dental landmarks (83.3%), 1 skeletal measurement (11.1%) and 2 dentoalveolar measurements (33.3%).

**Figure 2.**
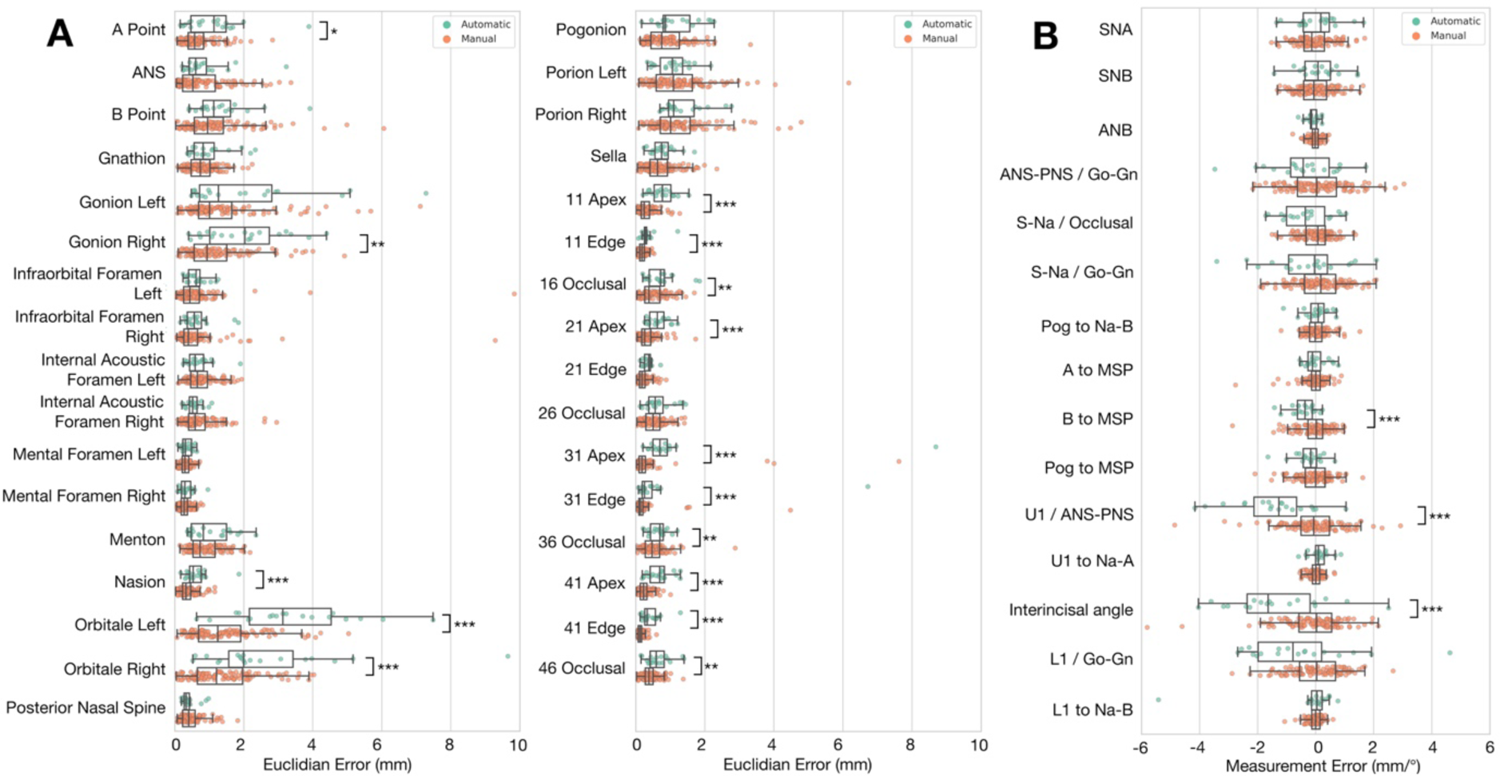
Localization and measurement error boxplots for automatic (green) and manual (orange) methods on 19 CT scans from the test set. (**A**) Localization errors (mm) for each landmark; (**B**) Measurement errors (mm/°) for each cephalometric variable. For each pair of results, statistically significant differences are indicated (* p<0.05; ** p<0.01; ***p<0.001).

### Three-Dimensional Visualization

We chose two subjects representative of our test dataset as well as the “outlier case” to illustrate our results. Figure 3 shows reference and predicted landmarks plotted on the fully automatically-obtained CT scan segmentations (Dot et al. 2022).

**Figure 3.**
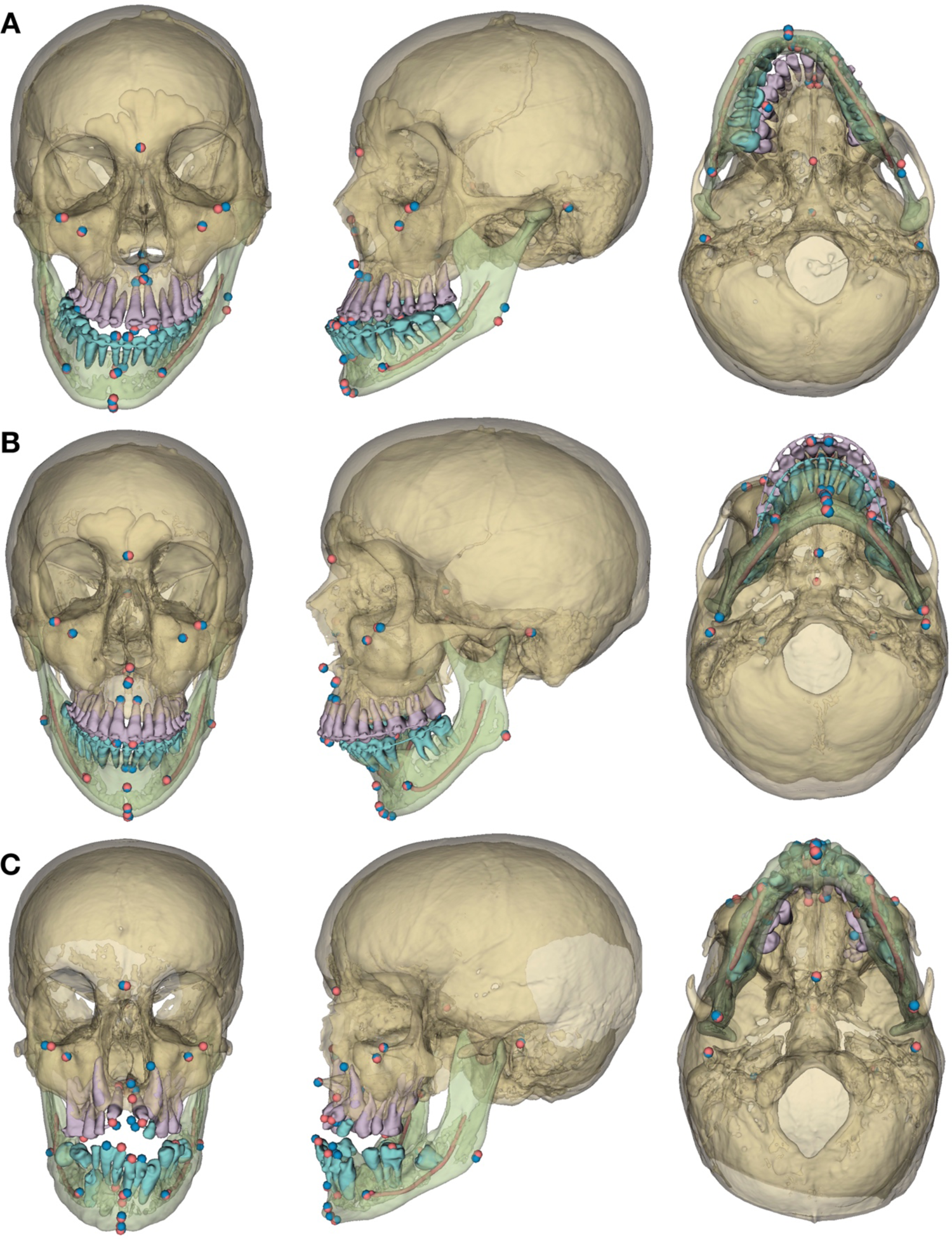
Frontal, ¾ left and inferior views of the 3D models, reference (red) and predicted (blue) landmarks for 3 subjects. (**A**) Prognathic and asymmetric mandible; (**B**) retrognathic mandible; (**C**) craniofacial syndrome “outlier case”, the errors in the predicted A point (at the level of the upper left canine apex) and the dental landmarks are to be noted.

## Discussion

The increasingly common use of 3D scans to assess complex maxillomandibular deformities and to plan orthognathic surgeries implies a critical need for clinical implementation of 3D cephalometric analyses. Such analyses currently require manual localization of 3D landmarks, a task that is time-consuming (± 15mn) and demands highly trained operators. In this study, we trained a DL network in order to localize 33 cephalometric landmarks automatically before evaluating the model on a challenging hold-out test set from clinical practice. The proposed DL pipeline took around one minute to localize the landmarks in a fully automatic manner. This amounts to a significant reduction of the time and effort needed for the task. The landmarks were localized with high accuracy, with 90.4% less than 2mm away from the manually localized reference landmarks.

Heterogeneity in the methods and datasets make studies reporting DL results notoriously difficult to evaluate and compare (Schwendicke, Singh, et al. 2021). The main strength of our study is that it provides a validation of our method based on a clinically relevant test dataset, randomly selected from a clinical sample of presurgical CT scans, 92.1% of which had metal artifacts. Moreover, we carefully constructed our reference test (manual landmarking) using the means of the six repetitions from a previously published R&R study for twenty of the test scans and asking one of this R&R study’s operators to label the 178 remaining scans. Overall, our results are comparable to current state-of-the art studies localizing landmarks on CBCT scans, some landmarks showing slightly better and other slightly worse localization results (Torosdagli et al. 2019; Zhang et al. 2020; Chen et al. 2021). However, previous studies lacked a clear definition of their dataset, localized fewer landmarks and evaluated their results following a cross-validation approach with no hold-out test dataset, which might question the generalizability of the results. It must be noted that our study focused on CT scans because it is the only imaging modality used for computer-assisted planning and personalized implant manufacturing for orthognathic surgery in our maxillofacial surgery department at this time. In future works we plan to use CBCT data in order to fine-tune our model and evaluate its accuracy on this other widespread imaging modality. Currently, our method does not perform automatic detection of the presence or absence of the landmarks; in the case of missing landmarks, those were deleted manually. We considered this approach sufficient, as it is easy for an operator to identify missing landmarks when running the cephalometric analysis, but other methods have been suggested to perform this task automatically (Lang et al. 2020; Chen et al. 2021).

The main goal of cephalometric landmarking is to perform linear and angular measurements which will ultimately provide clinical guidance. In order to evaluate the clinical usefulness of our DL-based method, our outcome set included lateral cephalometric measurements commonly found in R&R studies (van Bunningen et al. 2022) as well as three additional frontal measurements. We chose to perform 2D cephalometric measurements based on orthogonally projected landmarks because 3D cephalometric analysis remains complex, and thus beyond the scope of this study (Gateno et al. 2011). These measurements do not use the full potential of 3D cephalometry, but we believe they provide useful insight on the potential clinical usefulness of the method.

Concerning the skeletal landmarks, it has been shown that the reproducibility of manual landmarking was highly dependent on the type of landmark: landmarks localized on clear anatomical boundaries (*e*.*g*., sutures, spikes, holes) tend to be more reproducible than landmarks localized on skeletal contours (Sam et al. 2018). The comparison of our DL-based method with manual landmarking reproducibility shows that it is on par with trained clinicians for the localization of skeletal landmarks. Error-prone landmarks tend to be the same whether the landmarking is performed manually or automatically. Furthermore, even the landmarks with the worst accuracy results provided highly accurate cephalometric measurements or FH plane constructions, comparable with those obtained by clinicians. This confirms the need for evaluation outcomes other than MRE and SDR, as radial errors do not necessarily translate into clinically relevant errors (Gupta et al. 2016). Interestingly, landmarks localized on the craniofacial foramens showed excellent accuracy results, with 99.1% (*n* = 220) of the landmarks located within 2mm from the reference. These “novel” landmarks, which could not be localized on 2D cephalograms, could be used in future 3D cephalometric analyses (Naji et al. 2014; Lim et al. 2019; Dot et al. 2021).

Concerning the dental landmarks, despite good overall accuracy, the automated method provided less reliable results than the clinicians, with several automatic localizations showing errors statistically significantly larger than manual localization errors. The localization of these landmarks could probably be improved by refining their positions on the CT scan segmentation, for example using an additional knowledge-based method (Montúfar et al. 2018). When the patients’ intraoral scans are superimposed on the CT scans, for surgery planning for instance, they may also be segmented automatically and used for refining crown landmark localization (Hao et al. 2021 Nov 1).

We excluded one subject showing several landmarks with “very low” confidence levels, because such levels usually signal that the network did not work as expected and could lead to major errors. In this case, several landmarks (A Point and dental landmarks) showed errors >10mm (Appendix Table 7) and required operator corrections. These errors are probably due to the atypical anatomy of this subject, who exhibited a rare syndromic disease with several included teeth (Fig. 3C). From a clinical viewpoint, additional verification and correction of the results could be performed on a visualization of the predicted landmarks plotted on 3D models obtained fully automatically via DL (Fig. 3) (Wang et al. 2021; Dot et al. 2022).

To conclude, the proposed method achieved high accuracy on a test set of presurgical CT scans, providing results on par with those of clinicians for skeletal landmark localization and subsequent cephalometric measurements. The localization of dental landmarks still requires improvement to provide more reliable cephalometric measurements. Despite these promising results, our model requires additional testing in order to further evaluate its generalizability, reproducibility and robustness outside the scope of the present dataset. The data augmentation procedure that we applied during model training, based on image manipulations, should be helpful for the generalizability of the model (Shorten and Khoshgoftaar 2019) but it still has to be evaluated on an external test dataset including data from other clinical centers and CT machines. Afterwards, a prospective diagnostic efficacy study should evaluate the impact of using such an automated tool in routine clinical practice.

## Data Availability

All data produced in the present study are available upon reasonable request to the authors.

## Author Contributions

G. Dot contributed to the conception, design, data acquisition, analysis and interpretation, performed all statistical analyses, drafted and critically revised the manuscript. T. Schouman, P. Rouch and L. Gajny contributed to the conception, design and data interpretation, and critically revised the manuscript. S. Chang, F. Rafflenbeul and A. Kerbrat contributed to the data analysis and critically revised the manuscript. All authors gave final approval and agree to be accountable for all aspects of the work.

## Acknowledgments

The authors would like to thank C. Payer and all the team behind SpatialConfiguration-Net for sharing their research and codes.

## Ethical Approval

The IRB “Comité d’Ethique pour la Recherche en Imagerie Médicale” (CERIM) gave ethical approval for this research (number CRM-2001-051).

## Data availability

All data produced in the present study are available upon reasonable request to the authors.

