## Appendix for "Automatic Three-Dimensional Cephalometric Landmarking via Deep Learning"

*Supplemental Appendix*

### Materials and Methods

A DL pipeline was followed to localize cephalometric landmarks automatically on randomly selected craniomaxillofacial CT scans. We compared the results of our DL-based method on a hold-out test dataset (the index test) with those obtained by manual landmarking (the reference test). Our outcome set included localization accuracy, cephalometric measurements and comparison to manual landmarking reproducibility. The data were analyzed using R (http://www.r-project.org) and Python (http://www.python.org, version 3.7).

**Dataset**

Data were selected from a retrospective cohort of all consecutive patients having undergone orthognathic surgery in a single maxillofacial surgery department between January 2017 and December 2019, as described in a previous study (Dot et al. 2022). Patients referred to this center presented a wide variety of dentofacial deformities, came from various socioeconomic backgrounds, and were ethnically diverse. Patients were considered for inclusion whatever dental deformity they presented, with no minimum age. Exclusion criteria were refusal to participate in the research (all patients were contacted by mail). 200 subjects were randomly selected from this cohort, and 2 patients refused to participate. 198 subjects were eventually included in our dataset. The 198 CT scans performed before included subjects' orthognathic surgeries were de-identified and given an anonymization code (the anonymization chart was kept by the clinical investigator), no personal data was entered into the algorithm.

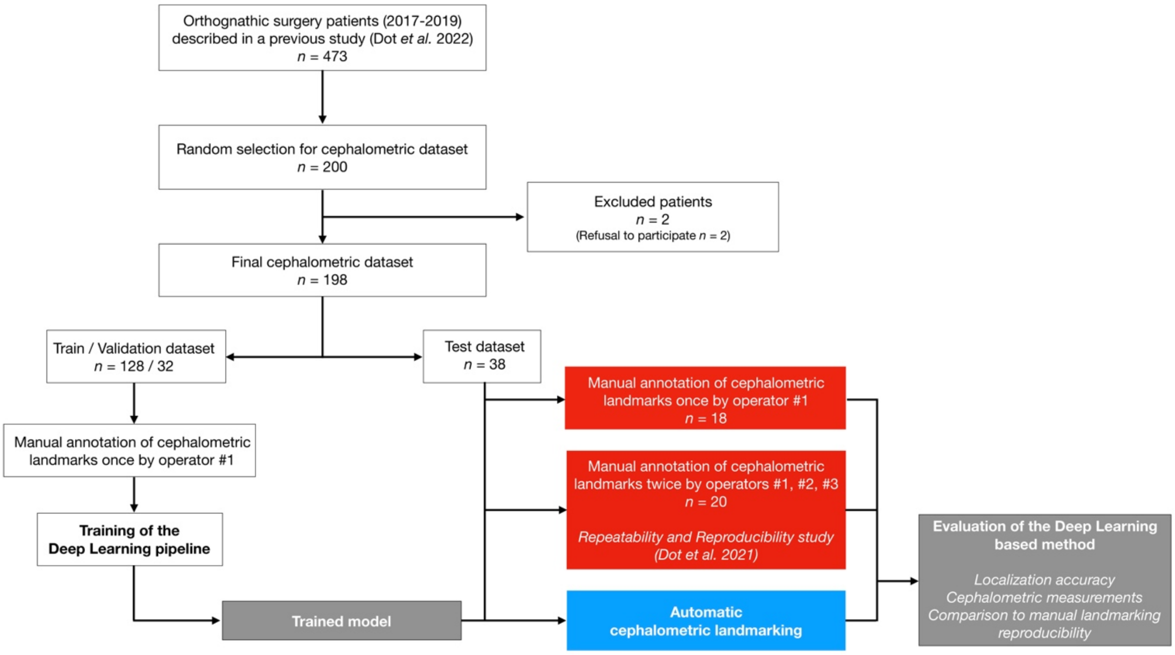

**Appendix Figure 1.** Data flow of patient selection, training and evaluation process. Red, reference test; blue, index test.

The vast majority of the included CT scans (*n* = 185, 93.4%) were acquired on a Discovery CT750 HD scanner (GE Healthcare, Chicago, USA) set at 100kVp, 50mAs, exposure time 730ms, slice thickness 0.625mm and slice increment 0.320mm. Field of view ranged from 165 to 320mm and pixel size ranged from 0.32 to 0.63mm. CT scans and patient characteristics are detailed in Appendix Table 1.

**Appendix Table 1.** CT scans and patient characteristics for the train/validation/test dataset.

| | Train | Validation | Test |
|---|---|---|---|
| Number of CT scans | 128 | 32 | 38 |
| Age, mean ± SD, years | 28 ± 11 | 28 ± 11 | 26 ± 8 |
| Gender, no. (%) | | | |
| Female | 76 (59.3) | 20 (62.5) | 20 (52.6) |
| Male | 52 (40.7) | 12 (37.5) | 18 (47.4) |
| Skeletal deformity, no. (%) | | | |
| Class I | 15 (11.7) | 3 (9.3) | 3 (7.9) |
| Class II[a] | 65 (50.8) | 17 (53.1) | 22 (57.9) |
| Class III[b] | 48 (37.5) | 12 (37.5) | 13 (34.2) |
| Syndromic deformity | 6 (4.7) | 2 (6.3) | 3 (7.9) |
| Metal artifacts, no. (%) | | | |
| Orthodontic materials | 104 (81.3) | 23 (71.9) | 33 (86.8) |
| Metallic dental filling/crown | 55 (43.0) | 14 (43.8) | 16 (42.1) |
| No metallic artifact | 14 (10.9) | 4 (12.5) | 3 (7.9) |
| Mean in-plane pixel size ($mm^2$) | 0.44 * 0.44 | 0.45 * 0.45 | 0.46 * 0.46 |
| Mean field of view (mm) | 228 | 229 | 233 |
| Mean slice thickness (mm) | 0.33 | 0.33 | 0.32 |
| Mean number of slices | 742 | 732 | 762 |
| Number of scans by CT Machine | | | |
| GEHC Discovery CT750 HD | 121 | 32 | 36 |
| GEHC Optima CT660 | 3 | | 1 |
| Other CT Machine[c] | 4 | | 1 |

[a]Prognathic maxilla and/or retrognathic mandible. [b]Retrognathic maxilla and/or prognathic mandible. [c]GEHC Revolution, Philips Ingenuity, Siemens SOMATOM. SD, Standard Deviation; GEHC: GE Healthcare.

#### Manual Landmarking (Reference Test)

Thirty-three commonly used landmarks, divided into skeletal (*n* = 21) and dental (*n* = 12), were manually annotated on each CT scan on the software Mimics (v.22.0, Materialise, Leuven, Belgium), following written and verbal instructions on the 3D description and annotation procedure for each landmark (please refer to (Dot et al. 2021) for more details about the written instructions provided to the operators). Definitions of the skeletal and dental landmarks are provided in Appendix Table 2 and 3, respectively.

**Appendix Table 2.** Definition of the "skeletal" landmarks localized in our study (L/R: Left/Right)

| Landmark name | Description |
|---|---|
| Nasion (Na) | Medial (and upper) point of the frontonasal suture |
| Orbitale L/R (Or-L / Or-R) | Lowest point of the orbital rim L/R |
| Anterior Nasal Spine (ANS) | Medial and most anterior point of the nasal spine |
| A Point (A) | Medial and most posterior point of the anterior concavity of the maxilla |
| B Point (B) | Medial and most posterior point of the anterior concavity of the mandible |
| Pogonion (Pog) | Medial and most anterior point of the mandible |
| Gnathion (Gn) | Medial and midpoint between Pog and Me |
| Menton (Me) | Medial and lowest point of the mandible |
| Gonion L/R (Go-L / Go-R) | Midpoint of the gonial angle L/R |
| Infraorbital Foramen L/R (IF-L / IF-R) | External & most distal point of the infraorbital foramen L/R |
| Internal Acoustic Foramen L/R (IAF-L / IAF-R) | External, most mesial and posterior point of the internal acoustic foramen L/R |
| Mental Foramen L/R (MF-L / MF-R) | External & most mesial point of the mental foramen L/R |
| Porion L/R (Po-L / Po-R) | External & uppermost point of the auditory canal L/R |
| Posterior Nasal Spine (PNS) | Medial & most distal point of the osseous palate |
| Sella (S) | Central point of the sella |

**Appendix Table 3.** Definition of the "dental" landmarks localized in our study (FDI World Dental Federation notation for teeth numbering)

| Landmark name | Description |
|---|---|
| 11, 21, 31, 41 edges (11E, 21E, 31E, 41E) | Midpoint of 11/21/31/41 incisal edges |
| 11, 21, 31, 41 apexes (11A, 21A, 31A, 41A) | Root apex of 11/21/31/41 |
| 16, 26 occlusal (16O, 26O) | Summit of the mesiopalatal cusp of 16/26 |
| 36, 46 occlusal (36O, 46O) | Central fossa of 36/46 |

All CT scans were manually annotated by the same operator (operator #1, a trained orthodontist with at least 5 years of clinical experience). 20 CT scans from the test set were manually annotated a second time by operator #1 and two times by operator #2 (a trained orthodontist with at least 5 years of clinical experience) and operator #3 (a final year postgraduate maxillofacial surgeon) following the procedure described in a previously published repeatability and reproducibility (R&R) study (Dot et al. 2021). The operators only had access to the CT data and segmentations, without any additional clinical information or pre-annotation, and had access neither to each other's results, nor to the index test results, nor to their first session results when performing the second session. Results were exported as an .xml file containing the x-, y-, z- coordinates of each landmark. The ground truth data used to train and test our deep learning model were the annotations of operator #1 for the CT scans landmarked once (*n* = 178), and the means of the 6 annotations by operators #1, #2, #3 for the CT scans landmarked several times (*n* = 20).

In the test set, some CT scans showed missing dental landmarks: 16O (*n* = 1), 26O (*n* = 1), 31A (*n* = 2), 31E (*n* = 2), 36O (*n* = 1), 46O (*n* = 2).

**Deep Learning-Based Landmarking (Index Test)**

All experiments were performed using the publicly-available SCN framework[1] running in Tensorflow v1.15.0 on our laboratory workstation (CPU AMD Ryzen 9 3900X 12-Core; 128 Gb RAM; GPU Nvidia Titan RTX 24Gb). All our trainings were performed on random image patches of size 128*128*192 voxels, mini-batch size of 1, learning rate $3.10^{-9}$ and momentum 0.99 (Nesterov's Accelerated Gradient) for 150,000 epochs. Data augmentation was performed on the fly using built-in methods detailed in (Payer et al. 2019). In order to predict each landmark coordinate, we detected the local heatmap maxima for each predicted volume heatmap. Accuracy metrics (distance between reference and predicted landmarks) on the validation set were used to select the final model. Based on the value of the local heatmap maxima, the confidence in a network prediction was considered "very low" when the value was below a threshold (0.4) established from the validation results.

Six networks were trained on our training set (*n* = 128) and evaluated on our validation set (*n* = 32):

- SCN#1 was trained on full scans with a "coarse" resolution of 0.90*0.90*0.62mm$^3$;
- SCN#2 to SCN#6 were trained on selected regions of interest (ROI) with a "fine" resolution of 0.45*0.45*0.31mm$^3$ (for orbitale, upper skull base, teeth & anterior maxilla and mandible regions) or 0.68*0.68*0.47mm$^3$ (for gonion region). These ROIs were created using the localization of the ground-truth landmarks and margins of 10mm (SCN#2 to SCN#5) or 30mm (SCN#6) in the -x, -y and -z directions.

[1] https://github.com/christianpayer/MedicalDataAugmentationTool-HeatmapRegression/tree/master/spine (accessed 2022 Jan 07)

Inference was performed on our test set ($n$ = 38) with a 2-stage method (Fig. 1B), following a sliding-window approach on image patches of the same size and resolution as the trained network. At stage 1, SCN#1 was used to predict the “coarse” localization of the landmarks. These results allowed us to extract the 5 ROIs using the localization of the predicted landmarks and margins of 10mm (SCN#2 to SCN#5) or 30mm (SCN#6) in the -x, -y and -z directions. At stage 2, SCN#2 to SCN#6 were used to predict the “fine” localization of the landmarks. Afterwards, the locally-predicted landmarks coordinates were transferred back into the general coordinate system for result evaluation. This method systematically localized 33 landmarks for each CT scan. In CT scans with missing landmarks (*i.e.* missing teeth), the corresponding predictions were considered as missing values and deleted by the operator.

Data preprocessing steps (e.g., normalization, rescaling, cropping of the image patches) were done automatically by the SCN. Other data processing steps (creation of ROIs, coordinate system transformations) were performed automatically using custom-made Python (http://www.python.org, version 3.7) scripts.

**Evaluation**

*Localization performance*

Each landmark was subjected to statistical analysis to compare the errors obtained from scans with reference constructed from 1 annotation with the errors obtained from scans with reference constructed from means of 6 annotations.

*Cephalometric measurements*

A conventional cephalometric analysis (Appendix Table 4) was conducted; nine 2D angles (degrees) and six 2D distances (mm) were calculated using orthogonal projections of the 3D landmarks on an automatically constructed midsagittal plane (MSP). MSP construction followed two steps: 1) the CT scans were segmented using a previously published DL-based automated method (Dot et al. 2022); 2) the MSP was computed thanks to the upper skull segmentation using a previously published automated method (Pinheiro et al. 2019). For each variable and each CT scan, the difference between the measurements obtained from reference landmarks and from predicted landmarks was computed and the proportion of measurements with differences under 2mm or 2° was calculated. Additionally, the accuracy of Frankfort horizontal (FH) plane construction (porion right/left and orbitale left) was evaluated by computing the absolute angular distances between reference and predicted FH planes.

**Appendix Table 4.** Skeletal and dentoalveolar cephalometric variables used in this study.

| Variable | Description |
|---|---|
| **Skeletal** | |
| SNA (°) | Angle between projected line S-Na and projected line Na-A (lateral view) |
| SNB (°) | Angle between projected line S-Na and projected line Na-B (lateral view) |
| ANB (°) | Angle between projected line Na-A and projected line Na-B (lateral view) |
| ANS-PNS / Go-Gn (°) | Angle between projected line ANS-PNS and projected line Mean_Go-Gn (lateral view) |
| S-Na / Go-Gn (°) | Angle between projected line S-Na and projected line Mean_Go-Gn (lateral view) |
| Pog to Na-B (mm) | Orthogonal distance between Pog and projected line Na-B (lateral view) |
| A to MSP (mm) | Orthogonal distance between A and projected MSP (frontal view) |
| B to MSP (mm) | Orthogonal distance between B and projected MSP (frontal view) |
| Pog to MSP (mm) | Orthogonal distance between Pog and projected MSP (frontal view) |
| **Dentoalveolar** | |
| S-Na / Occlusal plane (°) | Angle between projected line S-Na and projected occlusal plane (mean_16O_26O-mean_11E_21E) (lateral view) |
| U1 / ANS-PNS (°) | Angle between projected line mean_11E_21E-mean_11A_21A and projected line ANS-PNS (lateral view) |
| U1 to Na-A (mm) | Orthogonal distance between mean_11E_21E and projected line Na-A (lateral view) |
| Interincisal angle (°) | Angle between projected line mean_11E_21E-mean_11A_21A and projected line mean_31E_41E-mean_31A_41A (lateral view) |
| L1 / Go-Gn (°) | Angle between projected line mean_31E_41E-mean_31A_41A and projected line Go-Gn (lateral view) |
| L1 to Na-B (mm) | Orthogonal distance between mean_11E_21E and projected line Na-B (lateral view) |

*Comparison with manual landmarking reproducibility*

The results from a previous R&R study were used to assess the Bland-Altman 95% limits of agreement (LoA) of manual landmarking reproducibility (Dot et al. 2021). The proportion of predicted landmarks within these limits was computed for each *-x -y -z* axis. In addition, we applied ISO norm 5725 on the cephalometric variables to calculate the 95% LoA of manual measurement reproducibility (Appendix Table 5) (ISO 5725-2:2019). The proportion of predicted cephalometric variables within these limits was computed. For the CT scans included in the R&R study, statistical tests were used to compare automatic and manual results, and boxplots of the localization and measurement errors were computed.

**Appendix Table 5.** Bland-Altman 95% limits of agreement (LoA) of manual repeatability (2 repetitions) and reproducibility (3 operators) for the cephalometric variables (°/mm), calculated on 20 CT scans following the ISO 5725 standard.

| | **Repeatability 95% LoA** | **Reproducibility 95% LoA** |
|---|---|---|
| **Skeletal** | | |
| SNA (°) | 1.029 | 1.319 |
| SNB (°) | 0.975 | 1.318 |
| ANB (°) | 0.414 | 0.456 |
| ANS-PNS / Go-Gn (°) | 1.601 | 2.252 |
| S-Na / Go-Gn (°) | 1.621 | 1.979 |
| Pog to Na-B (mm) | 0.645 | 0.791 |
| A to MSP (mm) | 0.802 | 0.865 |
| B to MSP (mm) | 0.648 | 1.123 |
| Pog to MSP (mm) | 0.712 | 1.142 |
| **Dentoalveolar** | | |
| S-Na / Occlusal plane (°) | 0.826 | 1.107 |
| U1 / ANS-PNS (°) | 1.53 | 2.148 |
| U1 to Na-A (mm) | 0.405 | 0.464 |
| Interincisal angle (°) | 1.474 | 2.127 |
| L1 / Go-Gn (°) | 1.306 | 1.994 |
| L1 to Na-B (mm) | 0.38 | 0.504 |

#### Results

##### Localization performance

**Appendix Table 6.** Mean radial errors (mm), success detection rates (% (*n*)) and minimum/maximum radial error (mm) for each landmark on the test set with the outlier case included (*n* = 38). MRE, mean radial error; SD, standard deviation; Min., minimum radial error; Max., maximum radial error; L, left; R, right.

| | MRE ± SD | <2mm | <2.5mm | <3mm | Min. | Max. |
|---|---|---|---|---|---|---|
| 11 Apex | 1.7 ± 6.2 | 97.4 (37) | 97.4 (37) | 97.4 (37) | 0.2 | 39.0 |
| 11 Edge | 0.6 ± 1.0 | 97.4 (37) | 97.4 (37) | 97.4 (37) | 0.1 | 6.2 |
| 16 Occlusal | 1.3 ± 2.4 | 94.6 (35) | 94.6 (35) | 94.6 (35) | 0.1 | 11.2 |
| 21 Apex | 0.7 ± 0.4 | 100 (38) | 100 (38) | 100 (38) | 0.2 | 1.9 |
| 21 Edge | 0.7 ± 1.2 | 97.4 (37) | 97.4 (37) | 97.4 (37) | 0.1 | 7.8 |
| 26 Occlusal | 1.6 ± 3.5 | 91.9 (34) | 91.9 (34) | 91.9 (34) | 0.1 | 16.6 |
| 31 Apex | 1.5 ± 3.8 | 94.4 (34) | 94.4 (34) | 94.4 (34) | 0.2 | 22.2 |
| 31 Edge | 1.1 ± 3.3 | 91.7 (33) | 94.4 (34) | 94.4 (34) | 0.1 | 18.9 |
| 36 Occlusal | 1.8 ± 3.4 | 89.2 (33) | 89.2 (33) | 89.2 (33) | 0.2 | 12.7 |
| 41 Apex | 1.1 ± 2.9 | 97.4 (37) | 97.4 (37) | 97.4 (37) | 0.2 | 18.3 |
| 41 Edge | 0.6 ± 1.1 | 97.4 (37) | 97.4 (37) | 97.4 (37) | 0.1 | 7.1 |
| 46 Occlusal | 0.9 ± 1.8 | 97.2 (35) | 97.2 (35) | 97.2 (35) | 0.1 | 11.0 |
| A Point | 1.6 ± 2.9 | 86.9 (33) | 89.5 (34) | 89.5 (34) | 0.2 | 18.1 |
| Anterior Nasal Spine | 0.7 ± 0.7 | 94.7 (36) | 94.8 (36) | 97.4 (37) | 0.1 | 3.2 |
| B Point | 1.7 ± 1.5 | 65.8 (25) | 81.6 (31) | 92.1 (35) | 0.3 | 8.5 |
| Gnathion | 1.0 ± 0.6 | 92.1 (35) | 97.4 (37) | 100 (38) | 0.3 | 2.5 |
| Gonion L | 1.9 ± 1.7 | 68.4 (26) | 76.3 (29) | 86.8 (33) | 0.3 | 7.3 |
| Gonion R | 2.1 ± 1.4 | 50 (19) | 71.1 (27) | 73.7 (28) | 0.3 | 6.8 |
| Infraorbital Foramen L | 0.6 ± 0.3 | 100 (38) | 100 (38) | 100 (38) | 0.2 | 2.0 |
| Infraorbital Foramen R | 0.6 ± 0.5 | 97.4 (37) | 100 (38) | 100 (38) | 0.1 | 2.4 |
| Internal Acoustic Foramen L | 0.6 ± 0.4 | 100 (38) | 100 (38) | 100 (38) | 0.2 | 1.9 |
| Internal Acoustic Foramen R | 0.6 ± 0.6 | 97.4 (37) | 97.4 (37) | 97.4 (37) | 0.1 | 3.9 |
| Mental Foramen L | 0.4 ± 0.2 | 100 (38) | 100 (38) | 100 (38) | 0.1 | 0.8 |
| Mental Foramen R | 0.4 ± 0.3 | 100 (38) | 100 (38) | 100 (38) | 0.1 | 1.3 |
| Menton | 1.0 ± 0.6 | 94.7 (36) | 97.4 (37) | 100 (38) | 0.4 | 2.6 |
| Nasion | 0.7 ± 0.4 | 100 (38) | 100 (38) | 100 (38) | 0.1 | 1.9 |
| Orbitale L | 2.6 ± 2.0 | 44.7 (17) | 57.9 (22) | 68.4 (26) | 0.1 | 8.8 |
| Orbitale R | 2.6 ± 2.3 | 55.3 (21) | 65.8 (25) | 68.4 (26) | 0.3 | 9.7 |
| Pogonion | 1.1 ± 0.6 | 89.5 (34) | 97.4 (37) | 100 (38) | 0.2 | 3.0 |
| Porion L | 1.1 ± 0.5 | 89.5 (34) | 100 (38) | 100 (38) | 0.2 | 2.3 |
| Porion R | 1.3 ± 0.7 | 86.8 (33) | 89.5 (34) | 100 (38) | 0.3 | 2.8 |
| Posterior Nasal Spine | 0.5 ± 0.4 | 100 (38) | 100 (38) | 100 (38) | 0.1 | 1.5 |
| Sella | 0.8 ± 0.4 | 100 (38) | 100 (38) | 100 (38) | 0.2 | 2.0 |

**Appendix Table 7.** Radial errors (mm) for each landmark of the outlier case. L, left; R, right.

| | Euclidian Error (mm) |
|---|---|
| 11 Apex | 39.0 |
| 11 Edge | 6.2 |
| 16 Occlusal | 1.4 |
| 21 Apex | 1.3 |
| 21 Edge | 7.8 |
| 26 Occlusal | 16.6 |
| 31 Apex | 22.2 |
| 31 Edge | 18.9 |
| 36 Occlusal | 12.7 |
| 41 Apex | 18.3 |
| 41 Edge | 7.1 |
| 46 Occlusal | |
| A Point | 18.1 |
| Anterior Nasal Spine | 0.3 |
| B Point | 2.2 |
| Gnathion | 0.4 |
| Gonion L | 2.1 |
| Gonion R | 0.8 |
| Infraorbital Foramen L | 0.5 |
| Infraorbital Foramen R | 0.5 |
| Internal Acoustic Foramen L | 0.2 |
| Internal Acoustic Foramen R | 0.5 |
| Mental Foramen L | 0.5 |
| Mental Foramen R | 0.7 |
| Menton | 0.9 |
| Nasion | 1.5 |
| Orbitale L | 0.2 |
| Orbitale R | 3.6 |
| Pogonion | 1.0 |
| Porion L | 1.1 |
| Porion R | 0.8 |
| Posterior Nasal Spine | 0.5 |
| Sella | 0.9 |

**Appendix Table 8.** Mean radial errors (mm), success detection rates (% (*n*)) and minimum/maximum radial error (mm) for each landmark on the validation set (*n* = 32). MRE, mean radial error; SD, standard deviation; Min., minimum radial error; Max., maximum radial error; L, left; R, right.

| | MRE ± SD | <2mm | <2.5mm | <3mm | Min. | Max. |
|---|---|---|---|---|---|---|
| 11 Apex | 0.7 ± 0.3 | 100 (31) | 100 (31) | 100 (31) | 0.3 | 1.6 |
| 11 Edge | 0.5 ± 0.3 | 100 (31) | 100 (31) | 100 (31) | 0.1 | 1.2 |
| 16 Occlusal | 1.4 ± 2.3 | 83.9 (26) | 90.3 (28) | 90.3 (28) | 0.2 | 12.8 |
| 21 Apex | 0.7 ± 0.3 | 100 (30) | 100 (30) | 100 (30) | 0.2 | 1.3 |
| 21 Edge | 0.4 ± 0.2 | 100 (30) | 100 (30) | 100 (30) | 0.1 | 1.2 |
| 26 Occlusal | 1.3 ± 2.4 | 90 (27) | 93.3 (28) | 93.3 (28) | 0.3 | 13.4 |
| 31 Apex | 0.7 ± 0.4 | 100 (32) | 100 (32) | 100 (32) | 0.1 | 2.0 |
| 31 Edge | 0.5 ± 0.2 | 100 (32) | 100 (32) | 100 (32) | 0.2 | 1.0 |
| 36 Occlusal | 1.0 ± 1.9 | 96.4 (27) | 96.4 (27) | 96.4 (27) | 0.2 | 10.4 |
| 41 Apex | 0.7 ± 0.4 | 96.9 (31) | 96.9 (31) | 100 (32) | 0.2 | 2.6 |
| 41 Edge | 0.4 ± 0.2 | 100 (32) | 100 (32) | 100 (32) | 0.1 | 0.9 |
| 46 Occlusal | 1.2 ± 2.3 | 93.3 (28) | 96.7 (29) | 96.7 (29) | 0.2 | 13.1 |
| A Point | 1.1 ± 0.8 | 90.6 (29) | 93.8 (30) | 96.9 (31) | 0.3 | 4.0 |
| Anterior Nasal Spine | 0.9 ± 0.7 | 93.8 (30) | 93.8 (30) | 96.9 (31) | 0.1 | 3.3 |
| B Point | 1.9 ± 1.3 | 59.4 (19) | 68.8 (22) | 78.1 (25) | 0.3 | 5.0 |
| Gnathion | 1.0 ± 0.5 | 96.9 (31) | 100 (32) | 100 (32) | 0.2 | 2.3 |
| Gonion L | 1.4 ± 1.0 | 84.4 (27) | 90.6 (29) | 90.6 (29) | 0.2 | 4.8 |
| Gonion R | 1.5 ± 1.0 | 71.9 (23) | 78.1 (25) | 90.6 (29) | 0.2 | 3.8 |
| Infraorbital Foramen L | 0.8 ± 0.5 | 96.9 (31) | 100 (32) | 100 (32) | 0.1 | 2.3 |
| Infraorbital Foramen R | 0.9 ± 0.6 | 93.8 (30) | 96.9 (31) | 100 (32) | 0.2 | 2.8 |
| Internal Acoustic Foramen L | 0.5 ± 0.4 | 100 (32) | 100 (32) | 100 (32) | 0.2 | 1.6 |
| Internal Acoustic Foramen R | 0.7 ± 0.4 | 100 (32) | 100 (32) | 100 (32) | 0.1 | 1.9 |
| Mental Foramen L | 0.5 ± 0.5 | 96.9 (31) | 96.9 (31) | 100 (32) | 0.1 | 2.8 |
| Mental Foramen R | 0.4 ± 0.2 | 100 (32) | 100 (32) | 100 (32) | 0.1 | 1.0 |
| Menton | 1.1 ± 0.6 | 96.9 (31) | 96.9 (31) | 100 (32) | 0.4 | 2.9 |
| Nasion | 0.7 ± 0.4 | 100 (32) | 100 (32) | 100 (32) | 0.1 | 1.6 |
| Orbitale L | 1.8 ± 1.4 | 75 (24) | 78.1 (25) | 78.1 (25) | 0.2 | 5.1 |
| Orbitale R | 1.6 ± 1.1 | 75 (24) | 75 (24) | 90.6 (29) | 0.2 | 4.6 |
| Pogonion | 1.1 ± 0.6 | 93.8 (30) | 96.9 (31) | 100 (32) | 0.3 | 2.6 |
| Porion L | 1.1 ± 0.8 | 84.4 (27) | 90.6 (29) | 96.9 (31) | 0.3 | 3.1 |
| Porion R | 1.1 ± 0.6 | 90.6 (29) | 100 (32) | 100 (32) | 0.2 | 2.3 |
| Posterior Nasal Spine | 0.8 ± 1.2 | 93.8 (30) | 96.9 (31) | 96.9 (31) | 0.1 | 6.8 |
| Sella | 0.8 ± 0.4 | 100 (32) | 100 (32) | 100 (32) | 0.3 | 1.6 |

**Comparison with manual landmarking and measurement reproducibility**

**Appendix Table 9.** Proportion (% (*n*)) of predicted landmarks coordinates in the -x, -y and -z directions within Bland-Altman 95% limits of agreement of manual reproducibility (95% LoA) on the hold-out test set without the outlier case (*n* = 37). L, left; R, right.

| | **Within 95% LoA** | | |
|---|---|---|---|
| | X Axis | Y Axis | Z Axis |
| 11 Apex | 67.6 (25) | 67.6 (25) | 62.2 (23) |
| 11 Edge | 73 (27) | 81.1 (30) | 40.5 (15) |
| 16 Occlusal | 81.1 (30) | 91.9 (34) | 56.8 (21) |
| 21 Apex | 86.5 (32) | 75.7 (28) | 64.9 (24) |
| 21 Edge | 73 (27) | 83.8 (31) | 16.2 (6) |
| 26 Occlusal | 89.2 (33) | 86.5 (32) | 56.8 (21) |
| 31 Apex | 59.5 (22) | 27 (10) | 54.1 (20) |
| 31 Edge | 59.5 (22) | 64.9 (24) | 35.1 (13) |
| 36 Occlusal | 86.5 (32) | 89.2 (33) | 64.9 (24) |
| 41 Apex | 51.4 (19) | 64.9 (24) | 70.3 (26) |
| 41 Edge | 56.8 (21) | 56.8 (21) | 35.1 (13) |
| 46 Occlusal | 78.4 (29) | 81.1 (30) | 64.9 (24) |
| A Point | 100 (37) | 78.4 (29) | 83.8 (31) |
| Anterior Nasal Spine | 91.9 (34) | 91.9 (34) | 97.3 (36) |
| B Point | 91.9 (34) | 94.6 (35) | 91.9 (34) |
| Gnathion | 86.5 (32) | 89.2 (33) | 91.9 (34) |
| Gonion L | 89.2 (33) | 86.5 (32) | 83.8 (31) |
| Gonion R | 97.3 (36) | 70.3 (26) | 78.4 (29) |
| Infraorbital Foramen L | 97.3 (36) | 100 (37) | 100 (37) |
| Infraorbital Foramen R | 94.6 (35) | 94.6 (35) | 91.9 (34) |
| Internal Acoustic Foramen L | 97.3 (36) | 94.6 (35) | 100 (37) |
| Internal Acoustic Foramen R | 94.6 (35) | 97.3 (36) | 100 (37) |
| Mental Foramen L | 89.2 (33) | 89.2 (33) | 94.6 (35) |
| Mental Foramen R | 94.6 (35) | 86.5 (32) | 75.7 (28) |
| Menton | 89.2 (33) | 100 (37) | 91.9 (34) |
| Nasion | 83.8 (31) | 67.6 (25) | 94.6 (35) |
| Orbitale L | 75.7 (28) | 86.5 (32) | 86.5 (32) |
| Orbitale R | 73 (27) | 89.2 (33) | 86.5 (32) |
| Pogonion | 94.6 (35) | 89.2 (33) | 94.6 (35) |
| Porion L | 100 (37) | 83.8 (31) | 94.6 (35) |
| Porion R | 100 (37) | 94.6 (35) | 89.2 (33) |
| Posterior Nasal Spine | 97.3 (36) | 91.9 (34) | 91.9 (34) |
| Sella | 91.9 (34) | 91.9 (34) | 97.3 (36) |

**Appendix Table 10.** Proportion (% (*n*)) of predicted cephalometric measurements within Bland-Altman 95% limits of agreement of manual reproducibility (95% LoA) on the hold-out test set without the outlier case (*n* = 37).

| | **Within 95% LoA** |
|---|---|
| **Skeletal** | |
| SNA (°) | 86.5 (32) |
| SNB (°) | 91.9 (34) |
| ANB (°) | 91.9 (34) |
| ANS-PNS / Go-Gn (°) | 94.6 (35) |
| S-Na / Go-Gn (°) | 81.1 (30) |
| Pog to Na-B (mm) | 94.6 (35) |
| A to MSP (mm) | 100 (37) |
| B to MSP (mm) | 91.9 (34) |
| Pog to MSP (mm) | 94.6 (35) |
| **Dentoalveolar** | |
| S-Na / Occlusal plane (°) | 62.9 (22) |
| U1 / ANS-PNS (°) | 78.4 (29) |
| U1 to Na-A (mm) | 75.7 (28) |
| Interincisal angle (°) | 57.1 (20) |
| L1 / Go-Gn (°) | 77.1 (27) |
| L1 to Na-B (mm) | 78.4 (29) |

**Bland-Altman plots of landmarking localization.** For the 33 landmarks, the following plots show the deviations from the mean (blue line) of the 6 manual repetitions and the predictions for the 20 subjects included in the R&R study. Please note that the scales differ. Subject number 17 is the outlier case. Red lines show the ± 2×SD of reproducibility. SD, standard deviation.

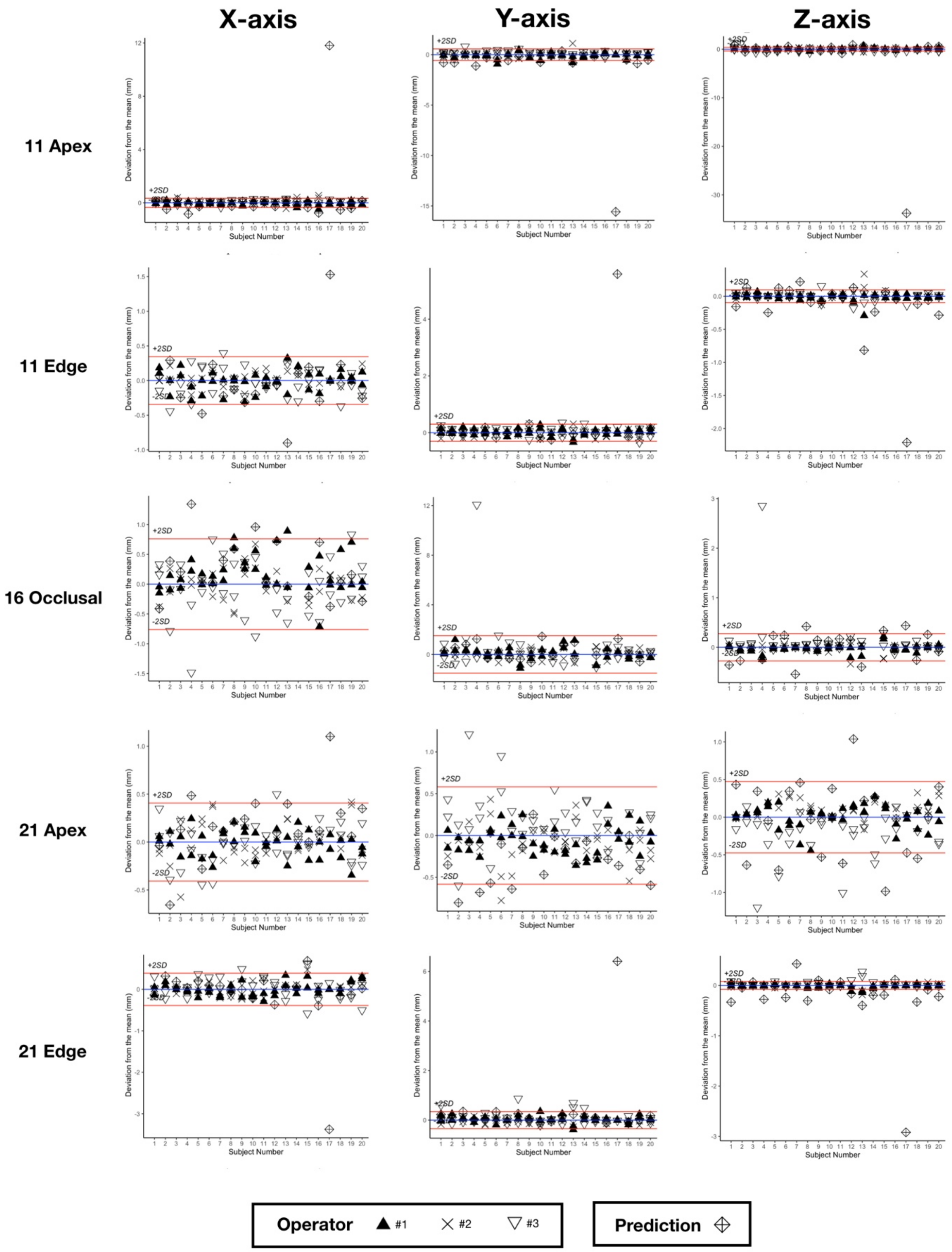

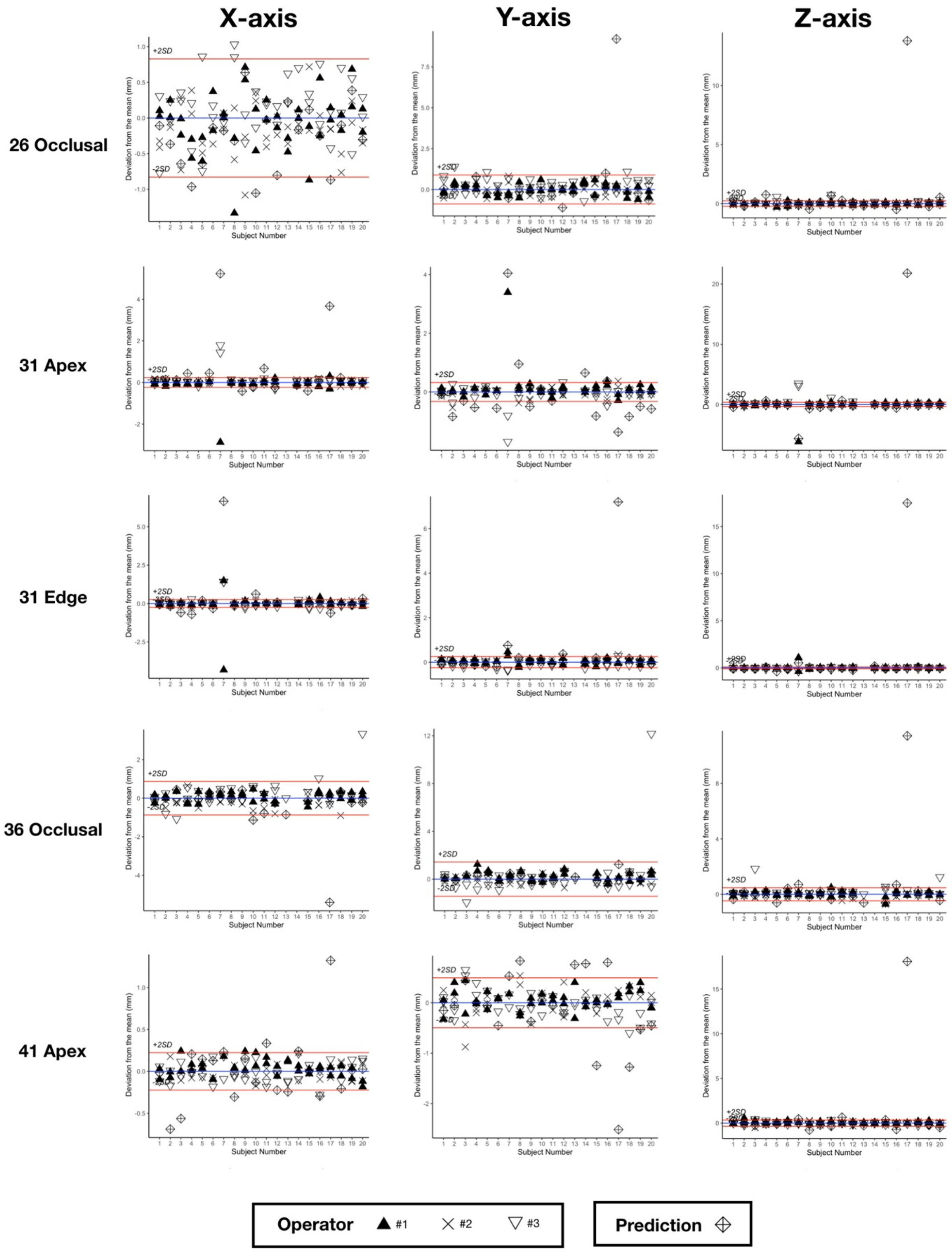
X-axis
Y-axis
Z-axis
26 Occlusal
31 Apex
31 Edge
36 Occlusal
41 Apex
Deviation from the mean (mm)
Subject Number
+2SD
-2SD
Operator #1 #2 #3
Prediction

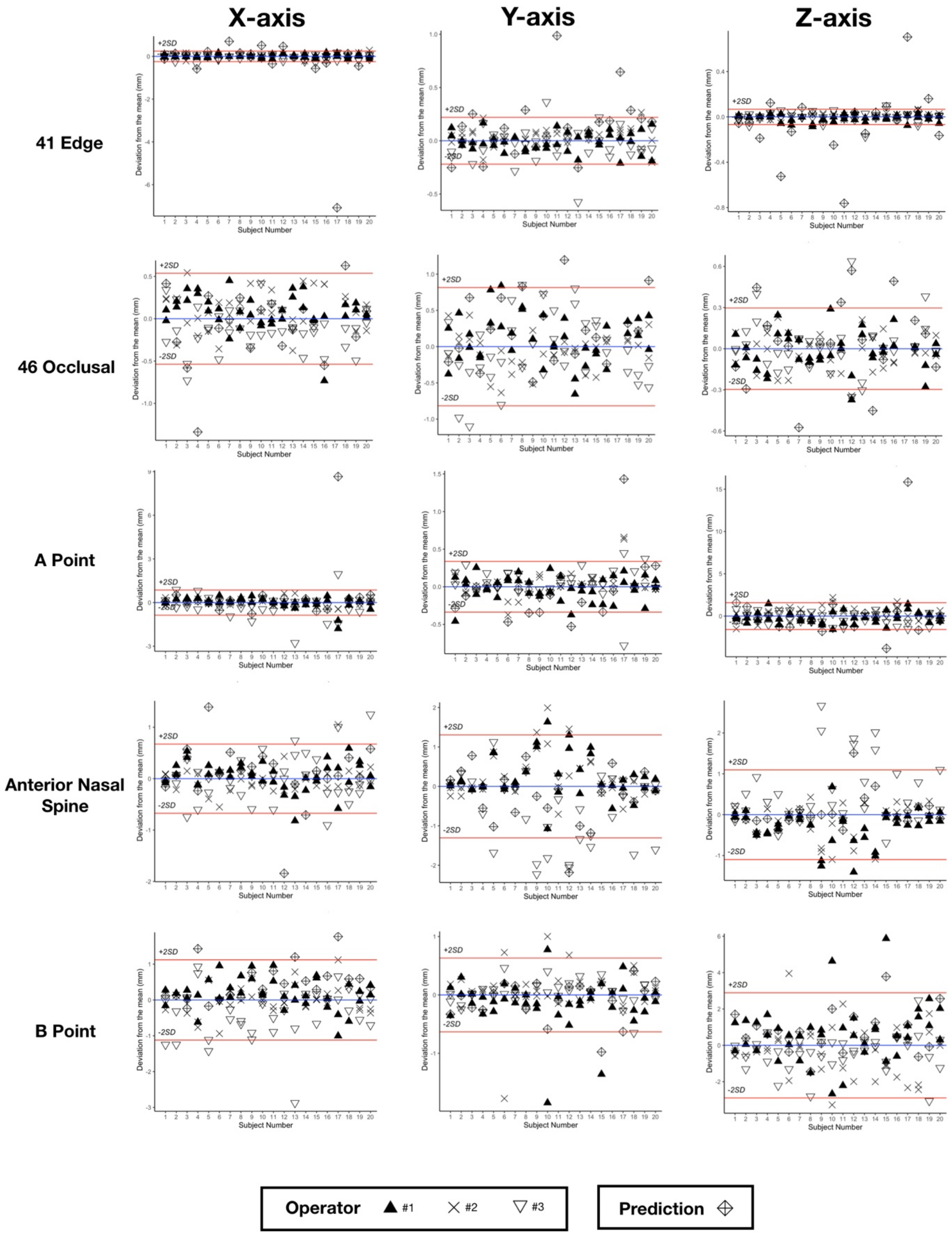

X-axis
Y-axis
Z-axis
41 Edge
46 Occlusal
A Point
Anterior Nasal Spine
B Point
Deviation from the mean (mm)
Subject Number
+2SD
-2SD
Operator #1 #2 #3
Prediction

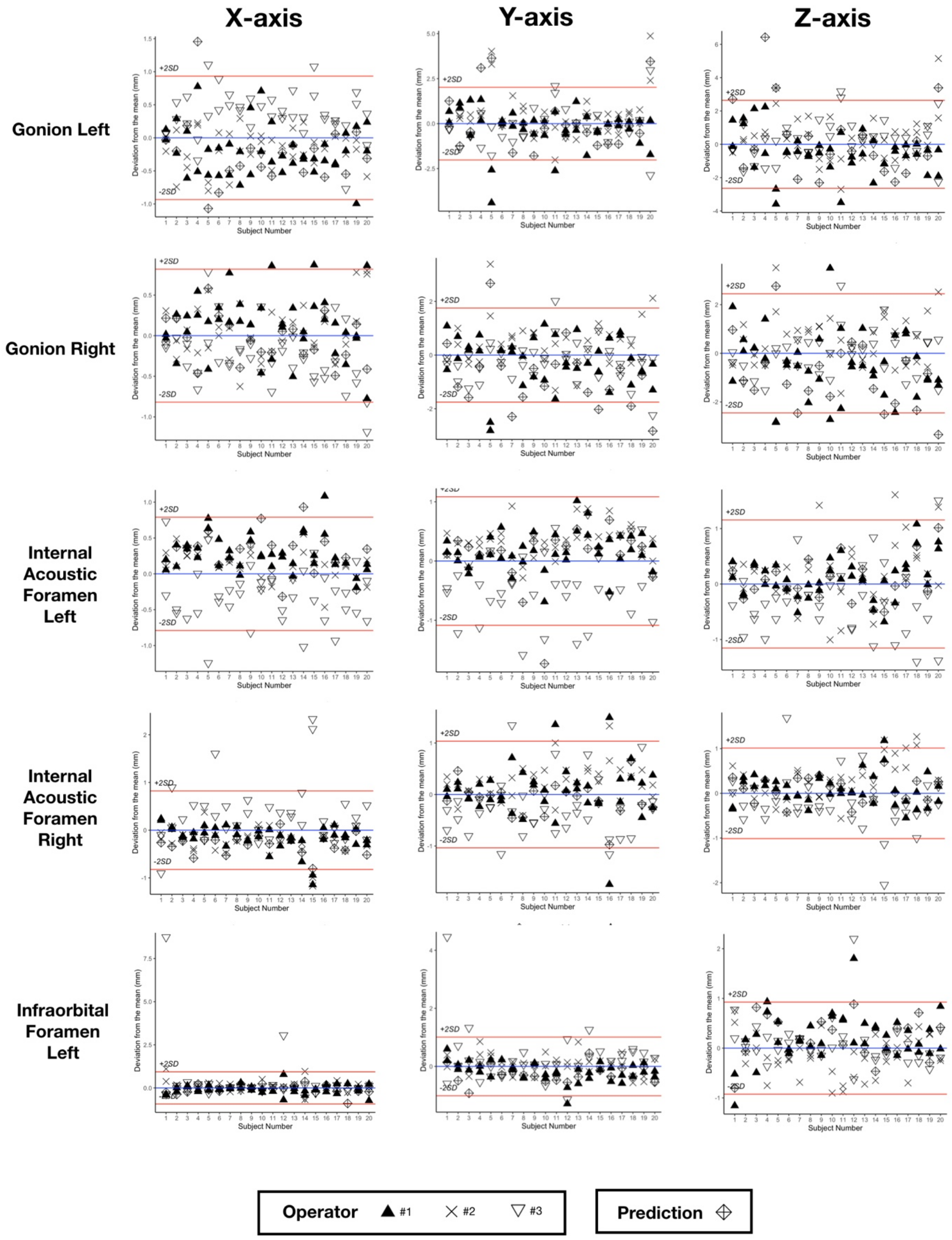

X-axis
Y-axis
Z-axis
Gonion Left
Gonion Right
Internal Acoustic Foramen Left
Internal Acoustic Foramen Right
Infraorbital Foramen Left
Deviation from the mean (mm)
Subject Number
+2SD
-2SD
Operator
#1
#2
#3
Prediction

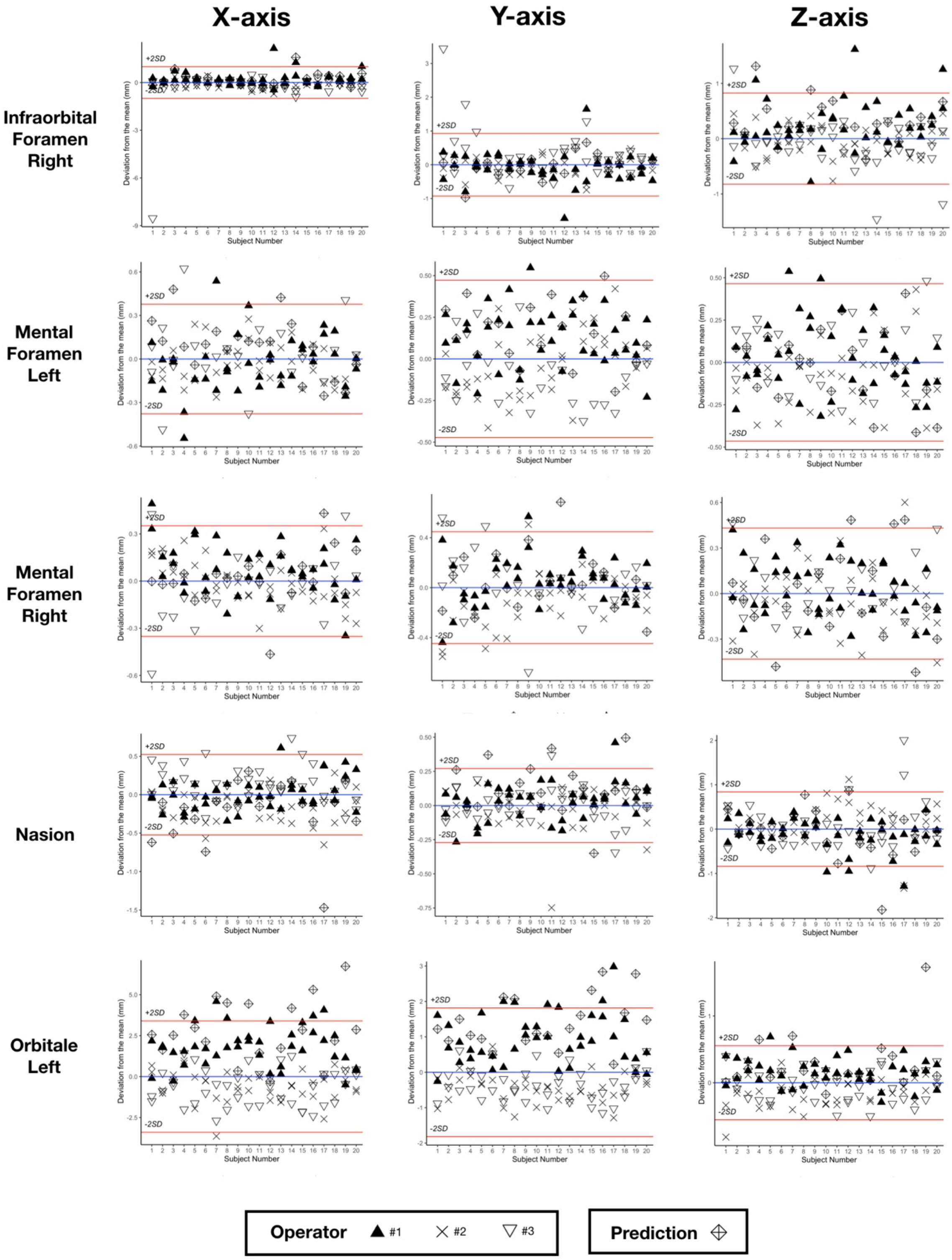
X-axis
Y-axis
Z-axis
Infraorbital Foramen Right
Mental Foramen Left
Mental Foramen Right
Nasion
Orbitale Left
Deviation from the mean (mm)
Subject Number
+2SD
-2SD
Operator #1 #2 #3
Prediction

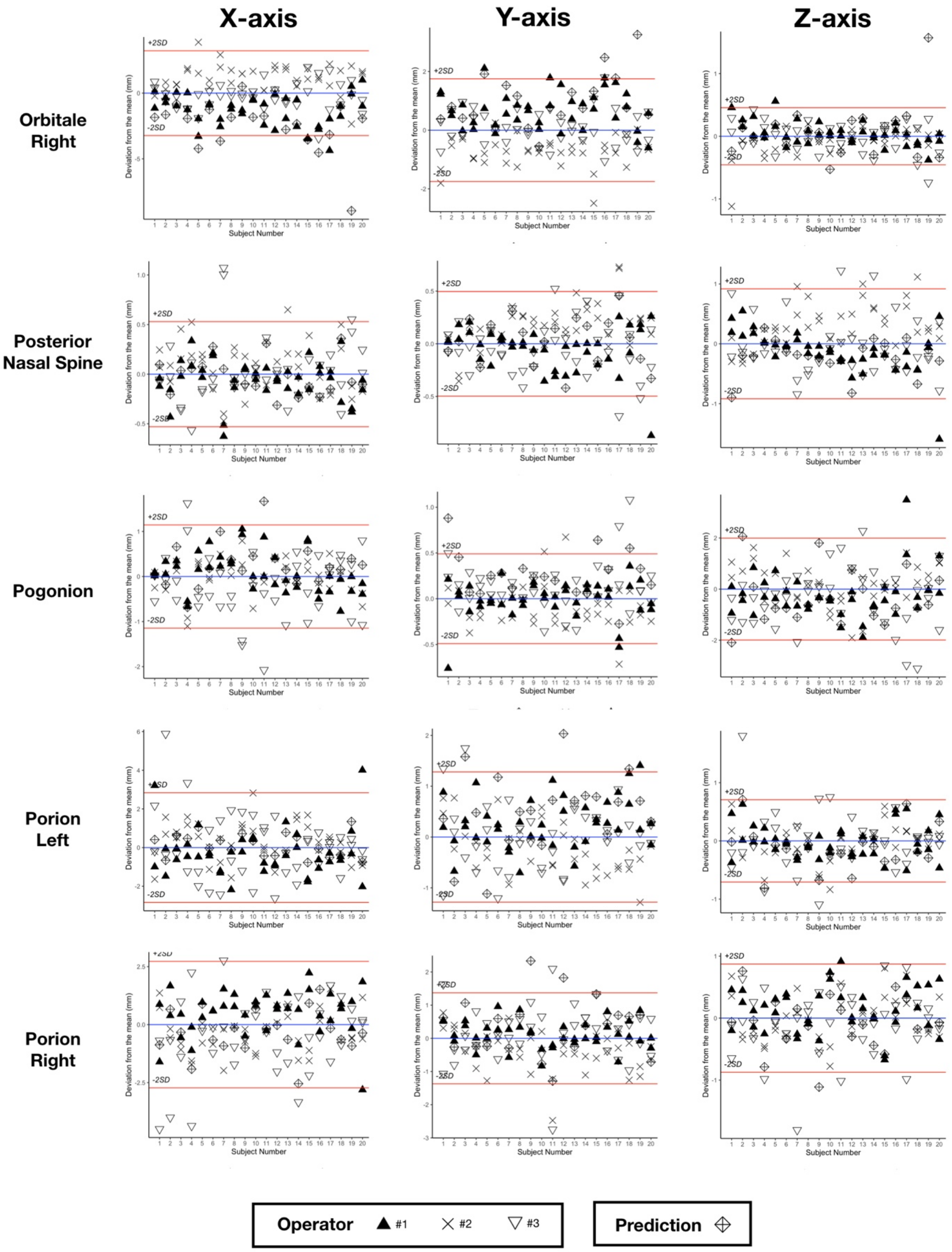

X-axis
Y-axis
Z-axis
Orbitale Right
Posterior Nasal Spine
Pogonion
Porion Left
Porion Right
Deviation from the mean (mm)
Subject Number
+2SD
-2SD
Operator #1 #2 #3
Prediction

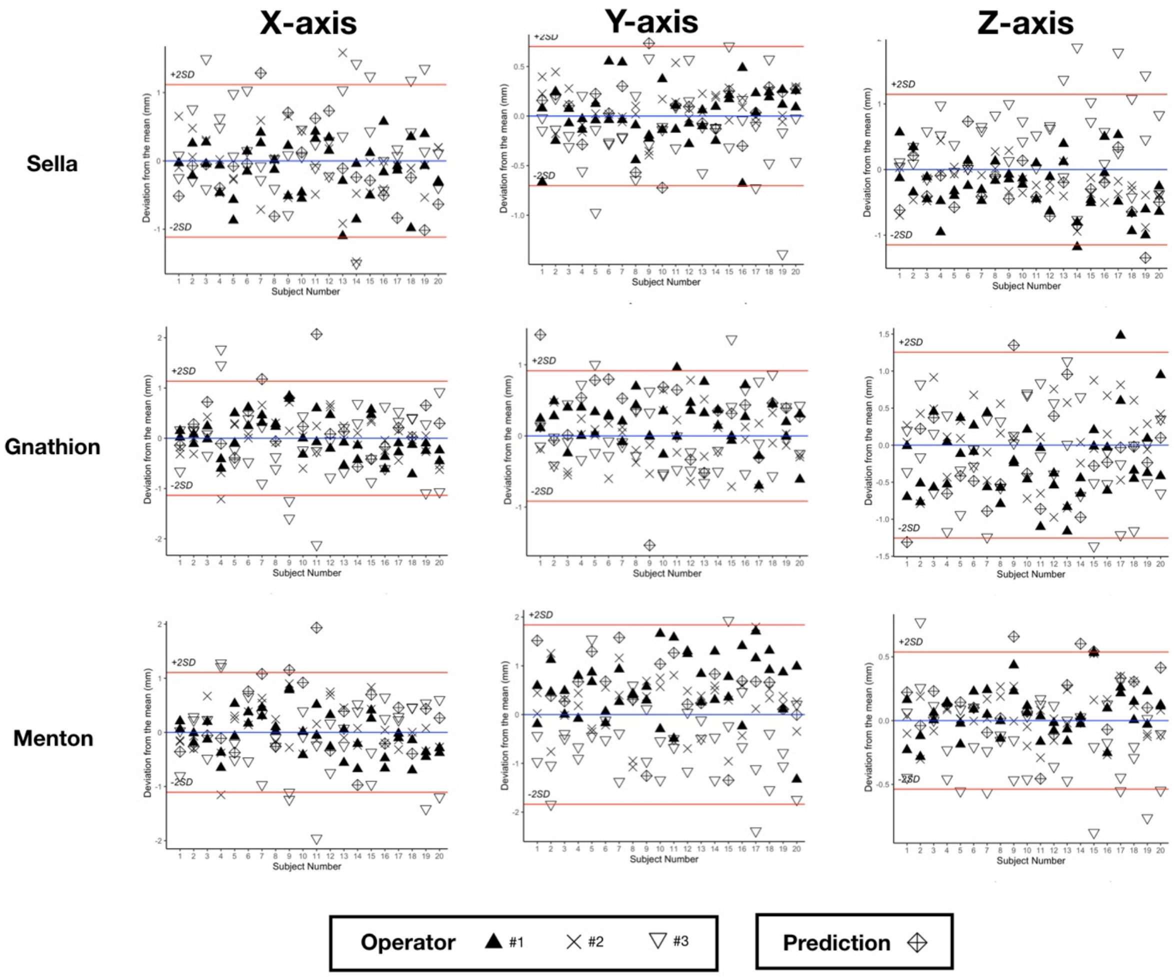

X-axis
Y-axis
Z-axis
Sella
Gnathion
Menton
+2SD
-2SD
Deviation from the mean (mm)
Subject Number
Operator
#1
#2
#3
Prediction

**Bland-Altman plots of cephalometric measurements.** For the 15 measurements, the following plots show the deviations from the mean (blue line) of the 6 manual repetitions and the predictions for the 20 subjects included in the R&R study. Please note that the scales differ. Subject number 17 is the outlier case. Red lines show the ± 2×SD of reproducibility. SD, standard deviation.

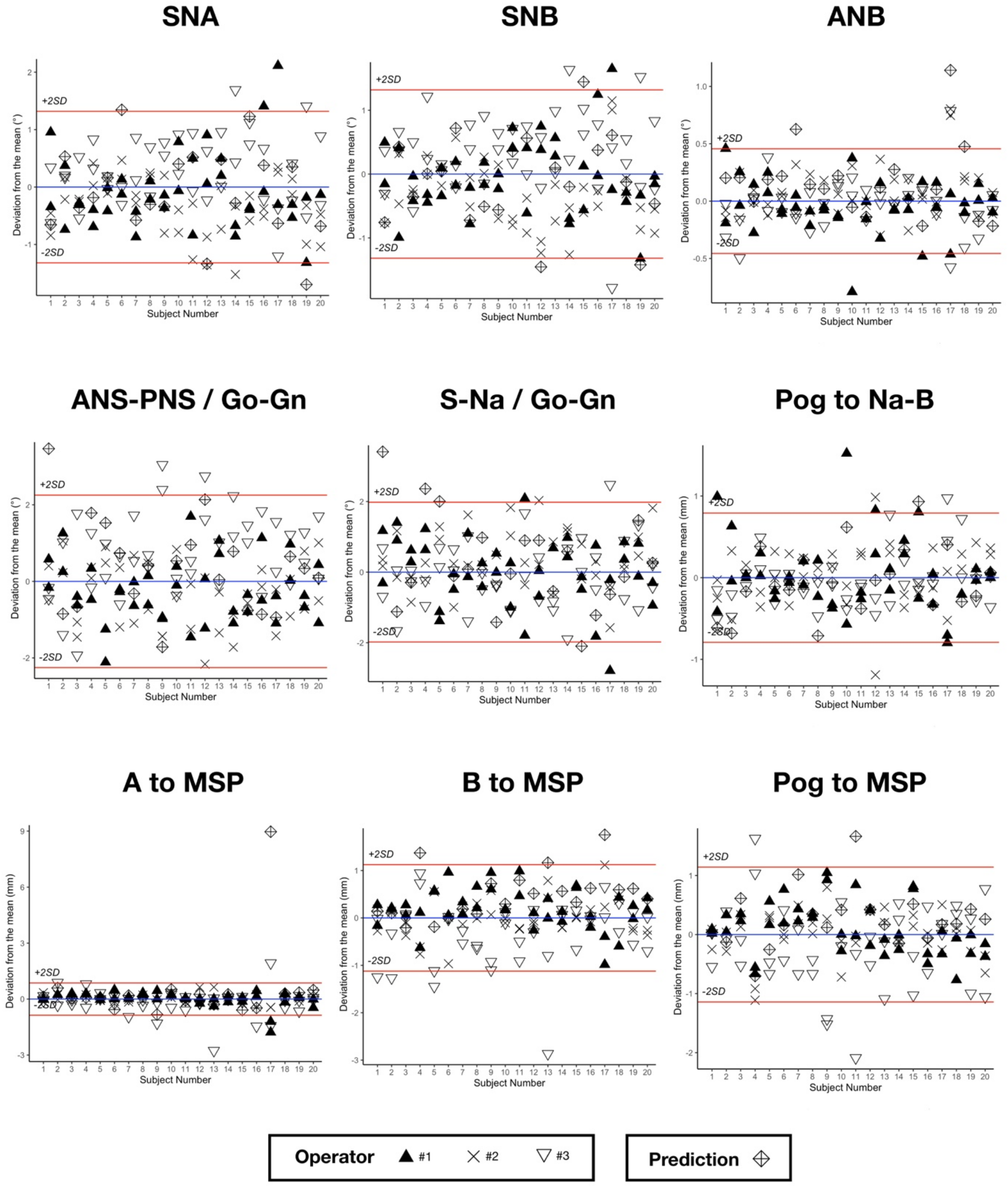

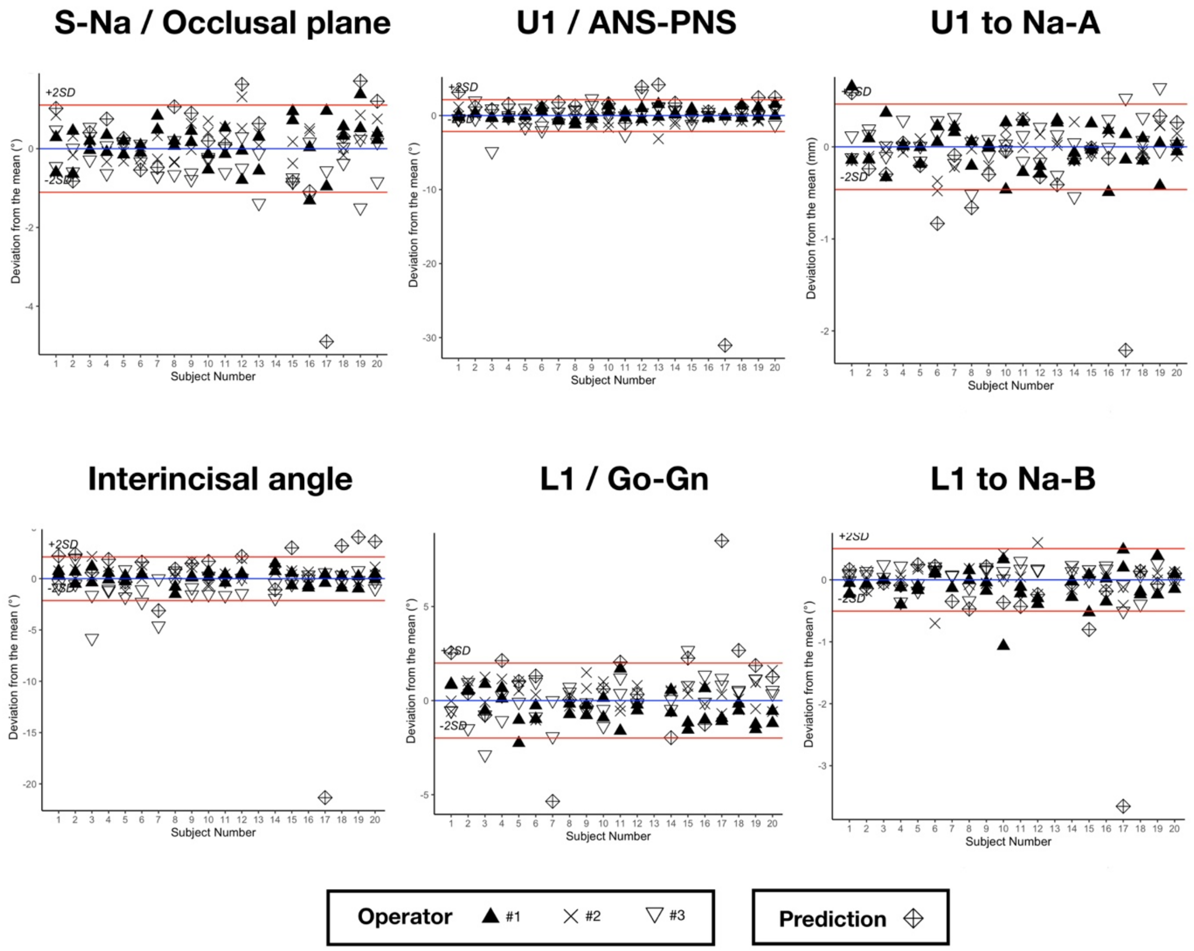
